# Polygenic and socioeconomic risk for high body mass index: 69 years of follow-up across life

**DOI:** 10.1101/2021.11.08.21265748

**Authors:** David Bann, Liam Wright, Rebecca Hardy, Dylan M Williams, Neil M Davies

## Abstract

**Background:** Genetic influences on body mass index (BMI) appear to markedly differ across life, yet existing research is equivocal and limited by a paucity of life course data. Better understanding changes across life in the determinants of BMI may inform etiology, the timing of preventative efforts, and the interpretation of increasing number of studies utilizing genetically-informed designs to study BMI. We thus used a birth cohort study to investigate differences in association and explained variance in the polygenic prediction of BMI from infancy to old age (2-69 years) in a single sample. A secondary aim was to investigate how a key purported environmental influence on BMI (childhood socioeconomic position) differed across life, and whether it operated independently and/or multiplicatively of genetic influences.

**Methods:** Data were from up to 2677 participants in the MRC National Survey of Health and Development, with measured weight and height from infancy to old age (12 timepoints from 2-69 years) and genetic data (obtained from blood samples at 53 years). We derived three polygenic indices derived from GWAS of a) adult BMI, b) recalled childhood body size, and c) childhood-adolescence BMI. We investigated associations of each polygenic index and BMI at each age and compared in terms of absolute effect size (β) and explained variance (R^2^). We used linear and quantile regression models, and finally investigated the additive or multiplicative role of childhood socioeconomic position.

**Results:** Mean BMI and its variance increased across adulthood. For polygenic liability to higher adult BMI (Khera et al), the trajectories of effect size (β) and explained variance (R^2^) diverged: explained variance peaked in early adulthood and plateaued thereafter, while absolute effect sizes increased throughout adulthood. For polygenic liability to higher childhood BMI, explained variance was largest in adolescence and early adulthood; effect sizes were marginally smaller in absolute terms from adolescence to adulthood. All polygenic indices were related to higher variation in BMI; effect sizes were sizably larger at the upper end of the BMI distribution. Socioeconomic and polygenic risk for higher BMI across life appear to operate additively; we found little evidence of interaction.

**Conclusion:** Our findings highlight the likely independent influences of polygenic and socioeconomic factors on BMI across life. Despite sizable associations, the BMI variance explained by each plateaued or declined across adulthood while BMI variance itself increased. This is suggestive of the increasing importance of chance (‘non-shared’) environmental influences on BMI across life.

## Introduction

Body mass index (BMI) is an important modifiable determinant of population health—its prevalence markedly increased from the 1980s onwards, and remains persistently high [1]. This drastic increase demonstrates the importance of environmental influences on BMI—population genetics do not change over such a short time span. Continuing evidence, however, has emerged on the link between genetic propensities and the level of BMI. For example, twin study estimates of heritability of BMI range from 47% to 90% [2] – with heritability typically highest in childhood. Polygenic indices in unrelated individuals predict approximately 8.5% of the variance in BMI [3].

Better understanding changes across life in the genetic determinants of BMI may inform etiology, the timing of preventative or weight loss efforts, and the interpretation of increasing number of studies utilizing genetically-informed designs to study BMI as either an exposure or outcome of interest [4–6]. Studies investigating genetic variation in the gene *FTO*—the first variant reliably linked to higher BMI—have repeatedly found that effect sizes are largest in earlier adulthood [7]. However, BMI is a complex and polygenic trait [3,5], necessitating a need to investigate how polygenic predictors of BMI differ across life.

Recent studies have used polygenic indices (also referred to as polygenic scores) to investigate associations with BMI at different life stages[4]. However, interpretation is hampered by a paucity of data across life on the same individuals. While samples of multiple birth cohorts can be used to approximate how associations differ by age, they may be confounded by the sizable cohort differences in links between polygenic indices and BMI [8]. Further, multiple polygenic indices [9] now exist for both childhood and adulthood BMI. Assessment of their performance in predicting phenotypic data requires replication using life course data to assess their use as plausible instrumental variables in Mendelian randomization analysis [3,5].

Other gaps in evidence motivate the need for future work. First, increases in BMI across life are marked by increases in its mean and its variance [10]. Conventional analytical approaches such as linear regression solely investigate mean differences. There is suggestive evidence that influence of genetic factors is strongest amongst those already higher in weight where health risks are greatest [11,12], yet this requires replication and formally testing.

Second, it is unclear how genetic and socioeconomic position (SEP) [13–15] influences on BMI operate together. While they may operate independently, it has been suggested that there may be multiplicative effects [16–18], such that genetic influences are largest amongst those in disadvantaged SEP whom have fewer resources available to protect against weight gain or to initiate/maintain weight loss. Alternatively, it has been hypothesized that genetic factors may confound associations between SEP and BMI [19] (or health outcomes more broadly [20]), or that in contrast to genetic influences shared environmental factors (such as of SEP) influence BMI in childhood but not adolescence [21].

We sought to address the above gaps in evidence using life course BMI data from a single national birth cohort study—this study, initiated in 1946, contains BMI data from infancy to old age. We used multiple polygenic indices, thought to indicate liability for either childhood or adult BMI. We investigated change across age in effect size and explained variance since each is likely to be informative; we also investigated the additive/multiplicative role of SEP and polygenic indices for BMI, and utilized quantile regression analysis to investigate associations across the BMI distribution.

## Methods

### Participants

The MRC National Survey of Health and Development [NSHD] (also known as the 1946 British birth cohort) is a longitudinal birth cohort study comprised of 5362 singleton births in mainland Britain born in a single week during March 1946 [22]. The cohort has been followed-up repeatedly across life, with blood samples obtained at 53 years and subsequently used for genotyping of common genome-wide genetic variation. DNA was extracted from whole blood samples obtained at 53 years, and purified using the Puregene DNA Isolation Kit (Flowgen, Leicestershire, UK) according to the manufacturer’s protocol. The study has received ethical approval from the North Thames Multicentre Research Ethics Committee (reference 98/2/121 and 07/H1008/168) and informed consent was provided.

### Measures

#### BMI

BMI (kg/m^2^) was derived from weight and height at 12 timepoints from 2-69 years of age (see Figure 1 for all ages); these were measured by health visitors, doctors, or nurses at all ages except 20 and 26 years where only self-reported data were available.

**Figure 1.**
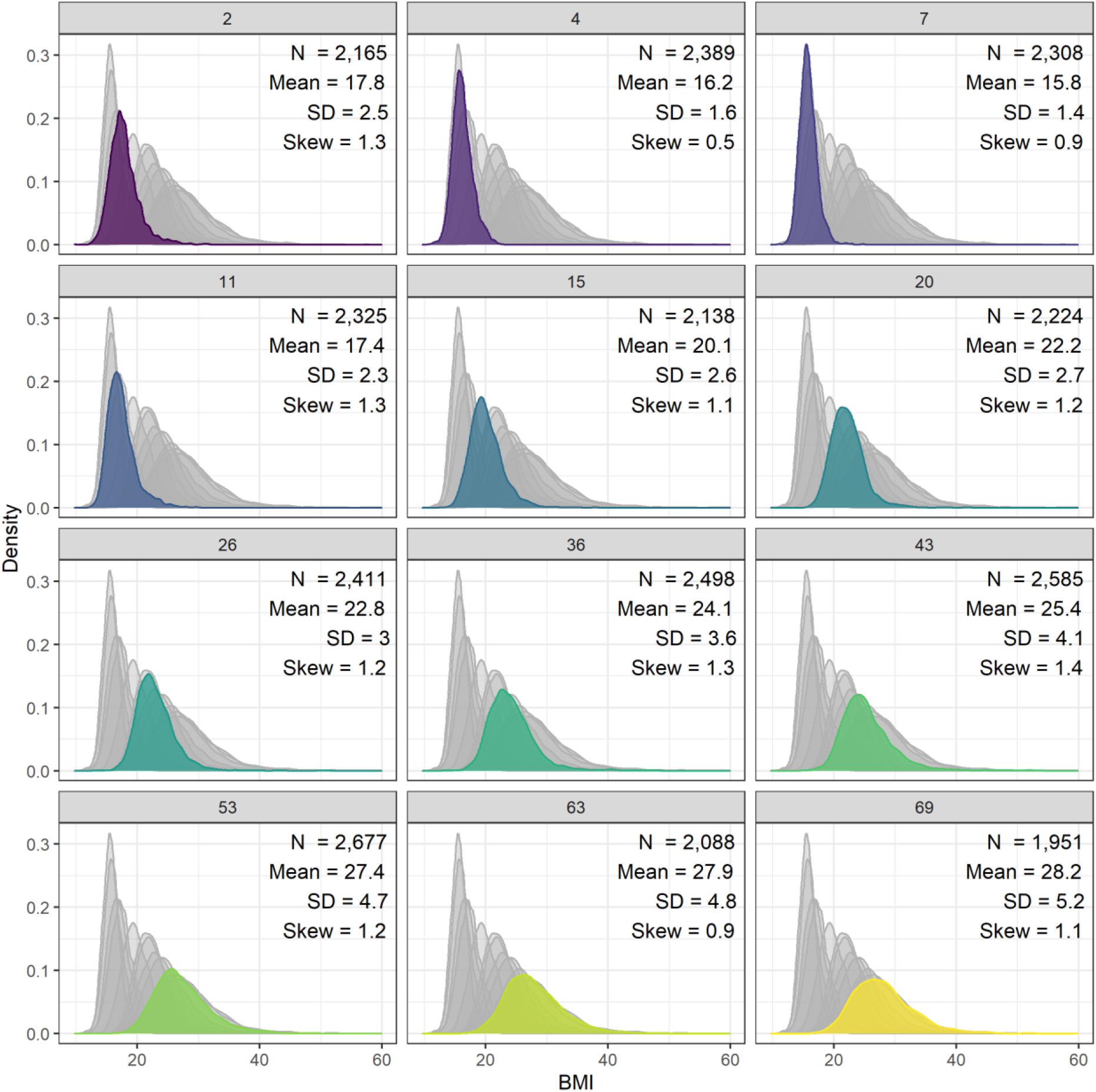
Histograms of body mass index from infancy to old age in the 1946 British birth cohort sample.

#### Polygenic indices

A total of 2851 individuals were genotyped using the DrugDev microarray (assaying 476,728 SNPs) platform. Quality control (QC) analyses performed using PLINK 1.9 [23]. Sample QC removed data on individuals with call rates <95%, extreme heterozygosity (µ ± 3 standard deviations), sex mismatches, relatedness and duplicates (>2), and principal component analysis (PCA) outliers. All participants were of European ancestry. Genotyped SNPs were excluded on the basis of the following parameters: call rate<95%, MAF<0.01 or HWE P<1e-4. The genotyped SNPs were used to impute information on missing common variants using the Haplotype Reference Consortium v1.1 reference panel, accessed via the Michigan Imputation Server [24]. QC of imputed data led to SNPs being excluded with INFO score<0.3 and MAF<0.005. Only biallelic SNPs were retained. Following these steps, data for 2794 individuals and 8,755,070 variants were retained for polygenic score calculations.

Three previously derived BMI-related polygenic scores were calculated for the NSHD participants:

i. An adult BMI score derived using UK Biobank data (N=119, 951), then tested in 4 cohorts from birth to adulthood; N=306,135 from Khera et al (2019) [3]. This yielded a score composed of 2,100,302 SNPs.
ii. A score for retrospectively reported childhood body size, derived from adults in UK Biobank self-reporting relative weight perception at age 10 (N=453,169) by Richardson et al (2020) [5]. This score consisted of 295 SNPs.
iii. A score for directly measured childhood BMI score derived from a meta-analysis of 41 childhood GWAS samples between ages 2 and 10 (N=61,111) from Vogelezang et al (2020) [6]. This yielded 25 SNPs for the score.

SNP data and weights for the adult BMI score and retrospective childhood body size score were downloaded from the PGS Catalog using trait PGS IDs PGS000027 and PGS000716, respectively [25]. Equivalent data for the direct childhood scores were extracted from the relevant publication and reformatted manually. To avoid strand ambiguities in each score, we removed palindromic SNPs from the two childhood scores (palindromic SNPs were already excluded by the authors of the adulthood BMI polygenic score prior to derivation). The three scores were then calculated from NSHD genotypes using Plink 2.0 [26], assuming additive genetic effects. Scores were based on 2,083,940 SNPs (99.2%) available from the original adult BMI score, 234 SNPs (79.3%) of the retrospective childhood body size score, and 21 SNPs (84.0%) of the direct childhood BMI score.

#### Socioeconomic position

To examine the association between BMI and childhood SEP and potential gene environment interactions between SEP and polygenic indices, we measured SEP as paternal occupational class at 4 years (Registrar General’s classification [RGSC] – I professional, II intermediate, III skilled non-manual, III skilled manual, IV semi-skilled, and V unskilled). To simplify interpretation, we converted this variable to a ridit score such that the resulting quantity in regression models shows the difference in BMI between lowest and highest SEP. To minimize missing data, we used information at 11 years for individuals missing SEP at age 4 (n = 125). As a robustness check, we alternatively measure SEP using maternal age at leaving education (10-23), again converted to a ridit score.

### Statistical analysis

First, we estimated the association of each polygenic index with BMI separately at each age (adjusted for sex) using linear regression. From these regressions, we extracted coefficients and incremental R^2^ values to examine the size of the association and the variance explained by age. We investigated associations on the absolute (kg/m^2^) and relative (percentage, percentile rank and standardized score) scales separately, since each may be informative; the absolute scale may aid comparability of effect sizes across adulthood as a 1 unit increase in BMI may have equivalent health risk; the relative scale may aid comparability across childhood and adulthood given sizable differences in mean BMI and its variation across age.

Second, we used quantile regression [27] to examine whether the association of BMI with polygenic indices differed across the distribution of BMI. Unlike linear regression, which estimates differences in the conditional mean of a distribution, quantile regression estimates differences in conditional quantiles of a distribution. Repeated across different quantiles, the method allowed us to examine differences in the shape (variability) and location (central tendency) of a distribution according to the values of an independent variable. We estimated quantile regressions at each decile (10th, 20th, …, 90th) for each polygenic index.

Third, we tested whether the relationship between SEP, polygenic indices and BMI was additive or multiplicative by regressing BMI on SEP, by including SEP × polygenic index interaction terms. We again repeated these regressions for absolute and standardized BMI indices, each polygenic index, and each measure of SEP (father’s social class and mother’s education).

Data cleaning and analysis was conducted using R version 4.1.0 [28]. We focused interpretation on estimates and measures of precision (95% CI) rather than binary interpretation of p-values [29].

### Sensitivity analyses

We first tested whether results differed by sex by conducting sex-stratified analyses. We then investigated if the associations were driven by weight and/or height – BMI (kg/m^2^) is a ratio measure and thus could reflect associations with height, particularly at younger ages. To account for this, we estimated separate associations with weight, height, and BMI; we also calculated a corrected weight-for-height index, dividing weight (kg) by height raised to a power that reduced the correlation between height and the index to zero at each age.

Our main analyses were conducted using those with observed BMI data at each age. Due to loss to follow-up, sample sizes differ across age. To explore whether this influenced our results, we 1) investigated whether polygenic indices were related to whether the participant had observed BMI at a given age; and 2) repeated analyses in samples with valid data for all follow-ups from a given age up to age 69, iterating across follow-ups (e.g., those followed from age 2 had complete case data at all timepoints, while those followed-up from 53 years had valid data from 53-69 years).

## Results

### Descriptive statistics

From age 7 onwards, BMI indices increased and exhibited more variability (higher SD); see Figure 1. All polygenic indices were moderately-strongly positively correlated. The adult index was correlated at r = 0.38-0.39 with both childhood indices, and childhood indices were correlated at 0.56 with each other. There was some evidence that polygenic indices differed by social class, indicating social patterning of genetic risk (Figure S1). Notably, participants from professional backgrounds have approximately 0.2 SD lower polygenic index values (for all indices) relative to sample averages.

### Polygenic index (adulthood) and BMI

The polygenic index for adult BMI derived from Khera et al. [3] was positively associated with BMI at all ages; Figure 2, top panel. The size of the association was small in infancy and childhood and increased in strength from early adolescence (age 11) to older adulthood (age 69). Effects sizes were largest at age 53 and remained similarly large at ages 63 and 69. A 1 SD increase in polygenic index was associated with 1.46 (95% CI: 1.26, 1.67) kg/m^2^ higher BMI at age 69. Findings were similar when examined in terms of percentage BMI (log transformed*100) differences (Figure S2). However, when examined in terms of standardized BMI differences (i.e., relative to the mean and SD at each age), effect sizes remained similar from ages 15-69 (Figure S2). This was likely due to the increasing variance of BMI across time, such that larger absolute (kg/m^2^) effects did not equate to bigger differences relative to the sample SD. Similarly, the incremental variance (R^2^) explained by the polygenic index peaked at age 26 and were slightly weaker thereafter—from 0.1 (95% CI: 0.08, 0.12) at age 26 to 0.08 (95% CI: 0.06, 0.1) at age 69 (Figure 2). These figure in mid-later adulthood are similar to those found by Khera et al (∼ 8.5%).

**Figure 2:**
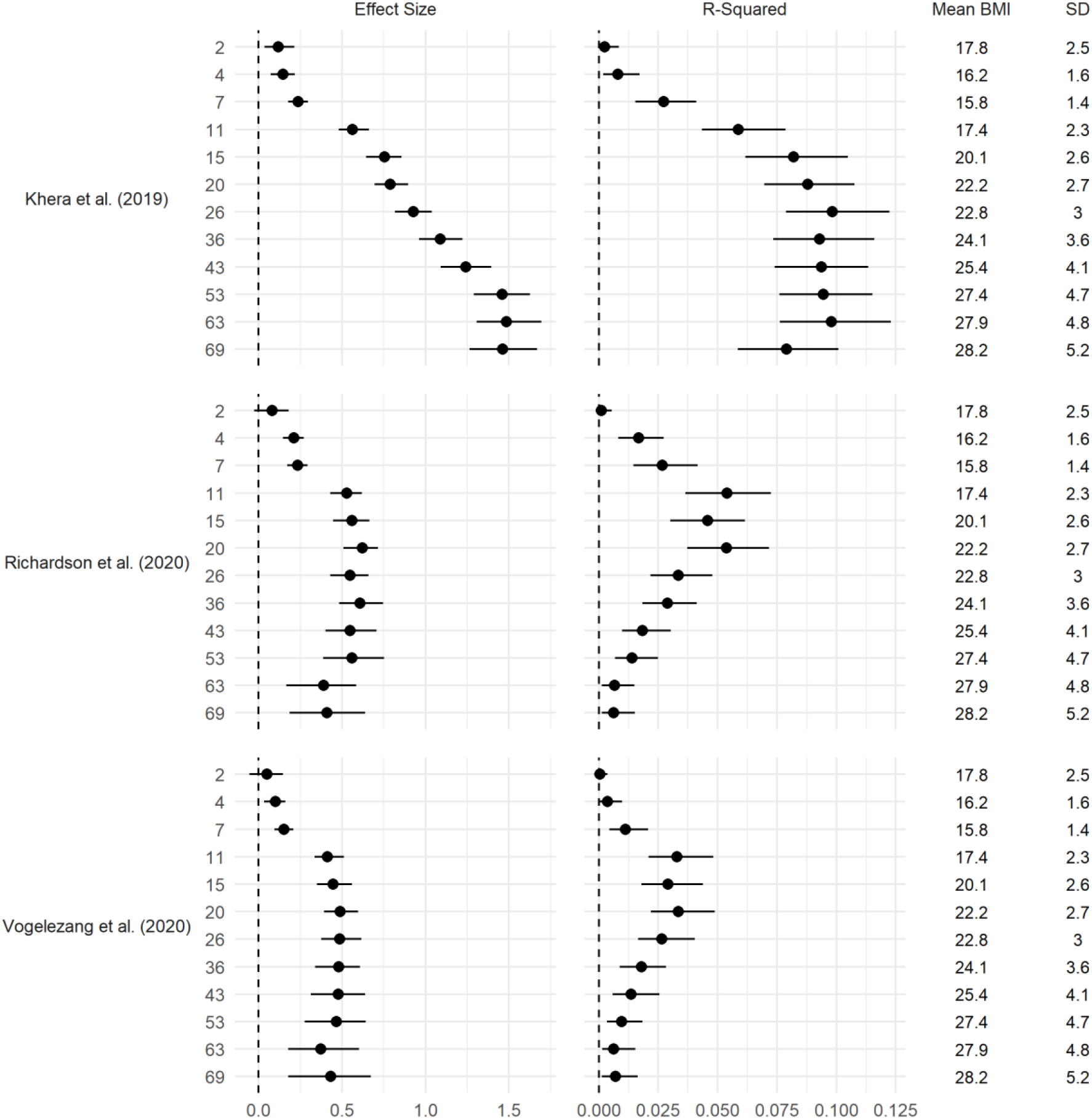
Association between polygenic indices and body mass index (BMI). Drawn from bivariate OLS regressions including adjustment for sex, repeated for each polygenic index and age at follow up. Left panel: coefficient difference in BMI per 1 SD increase in polygenic index (95% CI). Right panel: incremental R^2^ compared to OLS regression model of BMI on sex (95% CI estimated using bootstrapping [500 replications]).

The results of quantile regression analyses are shown in Figure 3. (Results with confidence intervals shown in Figure S3.) Associations between the polygenic index and BMI were progressively stronger at higher quantiles, suggesting that a higher polygenic index was associated with higher variability in BMI. For example, at age 69, the association between polygenic index and BMI was over twice as large at the 90th percentile (β = 1.93; 95% CI = 1.57, 2.30) as the 10th percentile (β = 0.87; 95% CI = 0.69, 1.20).

**Figure 3:**
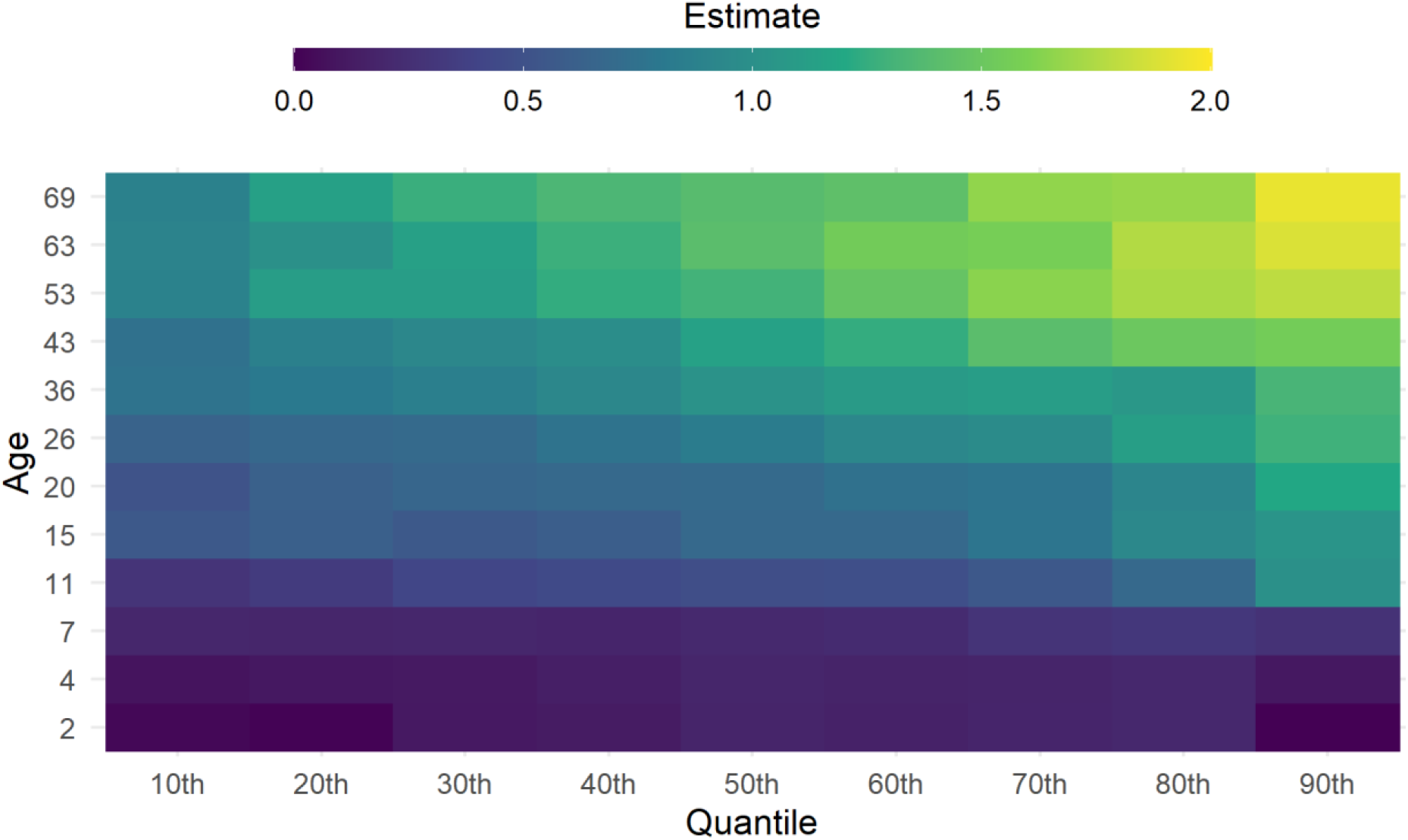
Heatmap of the association between Khera et al. [3] polygenic index and BMI. Drawn from quantile regressions including adjustment for sex, repeated at each follow up (y-axis) and decile (x-axis). The size of the coefficient is represented by a colour (see legend). Coefficients are interpreted analogously to linear regression: for example, Q50 shows the median (rather than mean) difference in body mass index per 1 SD increase in polygenic index.

### Polygenic index (childhood) and BMI

Results using the Richardson et al. [5] and Vogelezang et al. [6] polygenic indices for recalled childhood body size, and measured childhood BMI respectively are shown in Figure 2 (bottom two panels) and Figures S4-S5 (quantile regression results). The association between polygenic indices and BMI was largest in adolescence and early to mid-adulthood for both indices; associations were weak from ages 2-7, increased in size at age 11, and were marginally smaller at later ages (Figure 2). When examined on the relative scale (as percentage or z-score differences in BMI), the peak in effect size was more clearly evident from 11-20 (Figure S2), with declines in association thereafter corresponding to the increased sample BMI mean and SD with age. Similarly explained variance was highest in later adolescence to early adulthood (ages 11-20), and declined in mid to later adulthood (Figure 2). Overall, these polygenic indices explained less than 6% of variance in BMI at any age, which was less than that obtained when using the Khera et al. [3] polygenic index. Associations between the polygenic indices and BMI were progressively stronger at higher quantiles, particularly in adolescence and young-to-middle adulthood (Figures S4-S5).

### Polygenic indices and SEP interaction

More disadvantaged SEP in childhood (measured by father’s social class) was associated with higher BMI across multiple life stages; this association emerged from adolescence onwards and strengthened at each subsequent age, and was largely unchanged after adjustment for Khera et al. [3] polygenic indices (top panel, Figure 4). There was little evidence of SEP × polygenic index interaction; coefficients for interaction terms were close to zero at all ages with confidence intervals overlapping the null in almost all cases (bottom panel, Figure 4). Findings were similar across multiple specifications, including using childhood polygenic indices and measuring SEP with mother’s education level (Figures S6-S7). Further robustness checks using standardized or log BMI and SEP measured as mother’s education level measured on cardinal scale yielded qualitatively similar results (data available on request). The incremental explained variance (R^2^) attributable to SEP was low and similar across adulthood (< 2%, Figure S8).

**Figure 4:**
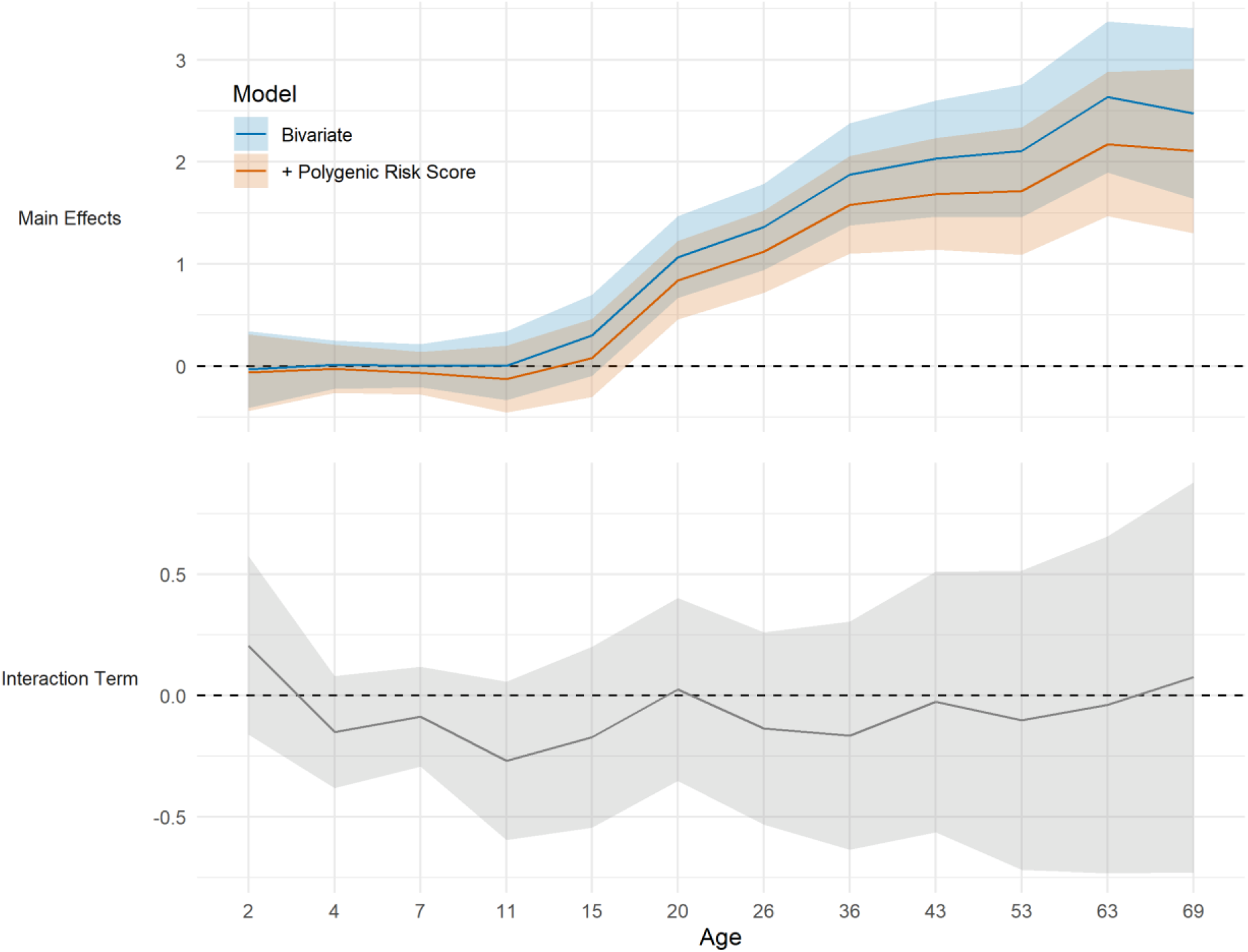
Association between childhood socioeconomic position and body mass index (BMI) across life. Top panel shows the kg/m^2^ difference in BMI in the lowest compared with highest socioeconomic position, before and after adjustment for Khera et al. [3] polygenic index for higher BMI. Bottom panel shows coefficients for the social class x polygenic index interaction term (null line is evidence for no interaction). SEP measured as father’s occupational class converted to ridit score. Results from top panel drawn from OLS regression models including adjustment for sex (blue solid line) and further adjustment for polygenic indices (orange dashed line). Results from bottom panel drawn from OLS regression models including adjusted for sex, polygenic index score [3] and SEP.

### Sensitivity analyses

Patterns of age difference in association between polygenic indices and BMI were similar in each sex (Figure S9), with some evidence that the associations were larger in females. Supplementary results suggested that differences in association across life were largely due to differences in weight rather than height (Figure S10); results were also similar when using weight-for-height indices constructed at each age using an optimal power of height, to remove the association between BMI and height (Figure S11).

Polygenic indices were related to missing BMI data at some ages (Figure S12). Notably, non-missing BMI data at ages 63 and 69 was related to lower than average Khera et al. [3] polygenic indices. Investigation of associations between polygenic indices and BMI using samples of the same participants across time showed broadly similar results as the main analysis (Figures S13-S18). However, there was some evidence that the plateauing of effect sizes in the Khera et al. [3] Index-BMI associations from 53 to 69 years was an artefact of differences in the samples at each age—when using the same sample across this age span, effect sizes were slightly higher at ages 63 and 69 (Figure S13).

## Discussion

### Summary of findings

Using life course BMI data spanning 2-69 years of age, and multiple polygenic indices for higher childhood and adulthood BMI, we found:

1. For polygenic liability derived from adult BMI (Khera et al. [3]), the trajectories of effect size and explained variance diverged across life: explained variance peaked in early adulthood and plateaued thereafter, while absolute effect sizes increased throughout adulthood.
2. For polygenic liability derived from recalled childhood body size and measured childhood BMI, explained variance was largest in adolescence and early adulthood; effect sizes were marginally smaller from adolescence to adulthood.
3. All polygenic indices were related to higher variation in BMI; effect estimates were sizable and larger at the upper end of the BMI distribution.
4. Childhood socioeconomic and polygenic risk for higher BMI across life appear to operate additively; with little evidence of interactions. The explained variance attributable to SEP on BMI was similar across adulthood.

### Comparison with previous studies and explanation of findings

Our findings are consistent with recent studies which used polygenic indices for higher adult BMI, typically in separate cohorts of different age spans. For example, Khera et al. [3] reported increasing effect sizes in regional cohorts followed-up from infancy to early adulthood (18 or 25 years). Sanz-de-Galdeano et al. [30] used separate cohorts and reported increasing strength of association from adolescence to early adulthood; while findings from a separate cohort suggested stability of effect sizes across older age. The results here show that these findings generalise to a single population born in 1946 and followed across the life course (2 to 69 years). Further research is required using multiple cohorts to examine how these results may differ by year of birth; the cohort used in this study was exposed to post-war rationing, and an increasingly obesogenic environment in midlife [31].

Our findings using polygenic indices for higher childhood BMI are also consistent with existing findings [5,6] – that indices derived using either recalled childhood weight or objectively measured childhood weight both have larger explained variance in childhood/adolescence/early adulthood. Our results suggest that such indices remain associated with higher BMI throughout early, mid and later adulthood, potentially leading to problems for their use in Mendelian randomization studies using childhood indices of BMI in relation to later outcomes.

Differences in findings across polygenic indices (or individual variants) suggests there may be age-specific mechanisms which link genetic liability to higher BMI. For example, genetic variants which have particularly stronger influence in early life may capture accelerated tempo of each life growth [32]. Multiple studies investigating *FTO* have reported largest effect sizes in early adulthood [7]: this was also found for the cohort used in this paper for *FTO* [33] and its nearby variants [32]. Further work is therefore required to elucidate the biological and behavioural mechanisms which link these polygenic indices to higher BMI.

Our findings highlight the importance of environmental influences on BMI across life. First, low childhood SEP was associated with higher adult BMI independently of polygenic risk; second, the mean and variance of BMI increased across life—this is likely to be due to environmental influences, since the explained variance attributable to genetic influence plateaued in early adulthood. The fact that explained variance attributable to SEP was small and remained similar across adulthood is suggestive of the increasing importance of chance or ‘non-shared’ environmental factors being increasingly important causes of between person in BMI variability across life. This finding is consistent with reported declines of heritability of BMI from adolescence to adulthood in twin studies [2]. It is notable that other traits have contrasting heritability patterns across life. For example, the heritability of cognitive performance appears to strengthen across life [34], potentially due to genetic influences indirectly influencing future environments which in turn strengthen genetic influence. In the context of BMI, such pathways may be sizably weaker relative to the large variability in the environment which influences BMI. Finally, all polygenic indices were associated with greater variability in BMI, with effect sizes largest in higher BMI centiles—one possible cause of this is the influence of unmeasured modifiers of association which may be environmental in origin [35].

### Strengths and Limitations

Strengths of this study include the use of life course data on a national birth cohort sample and use of multiple polygenic indices. Further, our analytical strategy enabled estimation of life course trajectories of effect size and explained variance; previous studies have tended to focus on either set of results yet both are informative. Our analysis also enabled formal testing of distributional effects, and the testing of the independent and/or multiplicative role of childhood SEP. Yet the necessary use of historic data had some inherent limitations. First, the cohort preceded the wider availability of body composition measures—thus, we cannot distinguish associations of fat or lean mass across the life course; it is possible that associations with these phenotypes may differ. Second, as in other prospective longitudinal studies missing data occurred. Genotyping occurred using blood samples measured at 53 years (in 1999); thus those with valid BMI data in early life yet no genotyping data were not included. However, we found little evidence that early life BMI was related to likelihood of having valid genotyping data at 53 years (Figure S18), though there was evidence that higher BMI during middle adulthood was related to having missing polygenic indices. Loss to follow-up occurred following genotyping from 53 to 69 years; the associations at later ages may have been downwardly biased (Figure S13).

## Conclusion

Our findings suggest sizable polygenic effects on BMI which differ in terms of size of association and explained variance across life. Findings also highlight the importance of the environment—adverse early life SEP was associated with higher BMI independently of polygenic risk, and increases in the population mean and variability of BMI across adulthood lead to stability of explained variance despite increasing effect sizes.

## Data Availability

Data are available via application from the LHA study team.

https://www.nshd.mrc.ac.uk/data

## Statements

### Author contributions

DB, NMD and LW conceived and designed the study. DW derived the polygenic indices. LW analysed the data. DB wrote the first draft and conducted initial analyses. All authors provided critical revisions and all authors read and approved the submitted manuscript.

### Data availability

Data are available via application from the following: https://www.nshd.mrc.ac.uk/data. The code used to run the analysis is available at https://osf.io/psbvy/.

