## supplemental file for "Polygenic and socioeconomic risk for high body mass index: 69 years of follow-up across life"

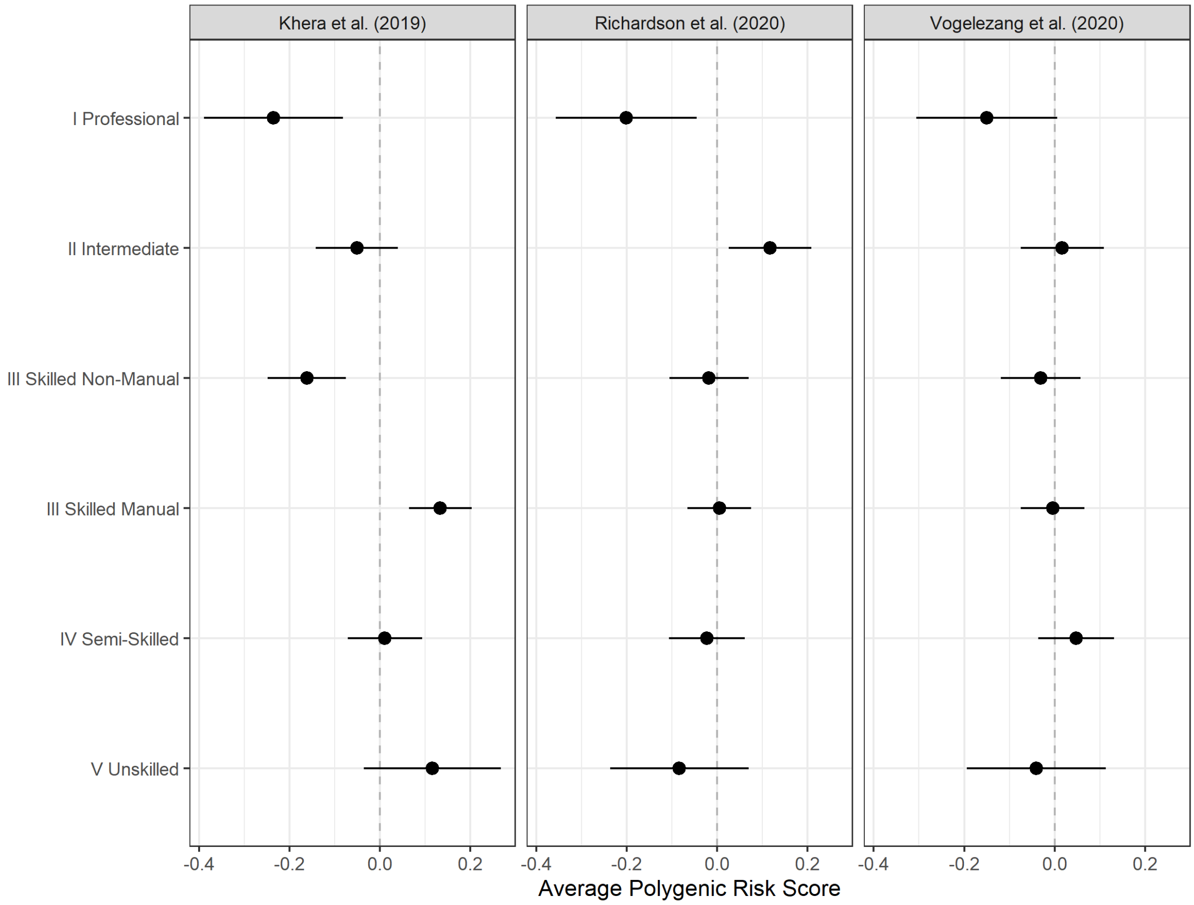


*Figure S1: Average polygenic risk scores by father's occupational class at age 4 (+ 95% confidence intervals).*

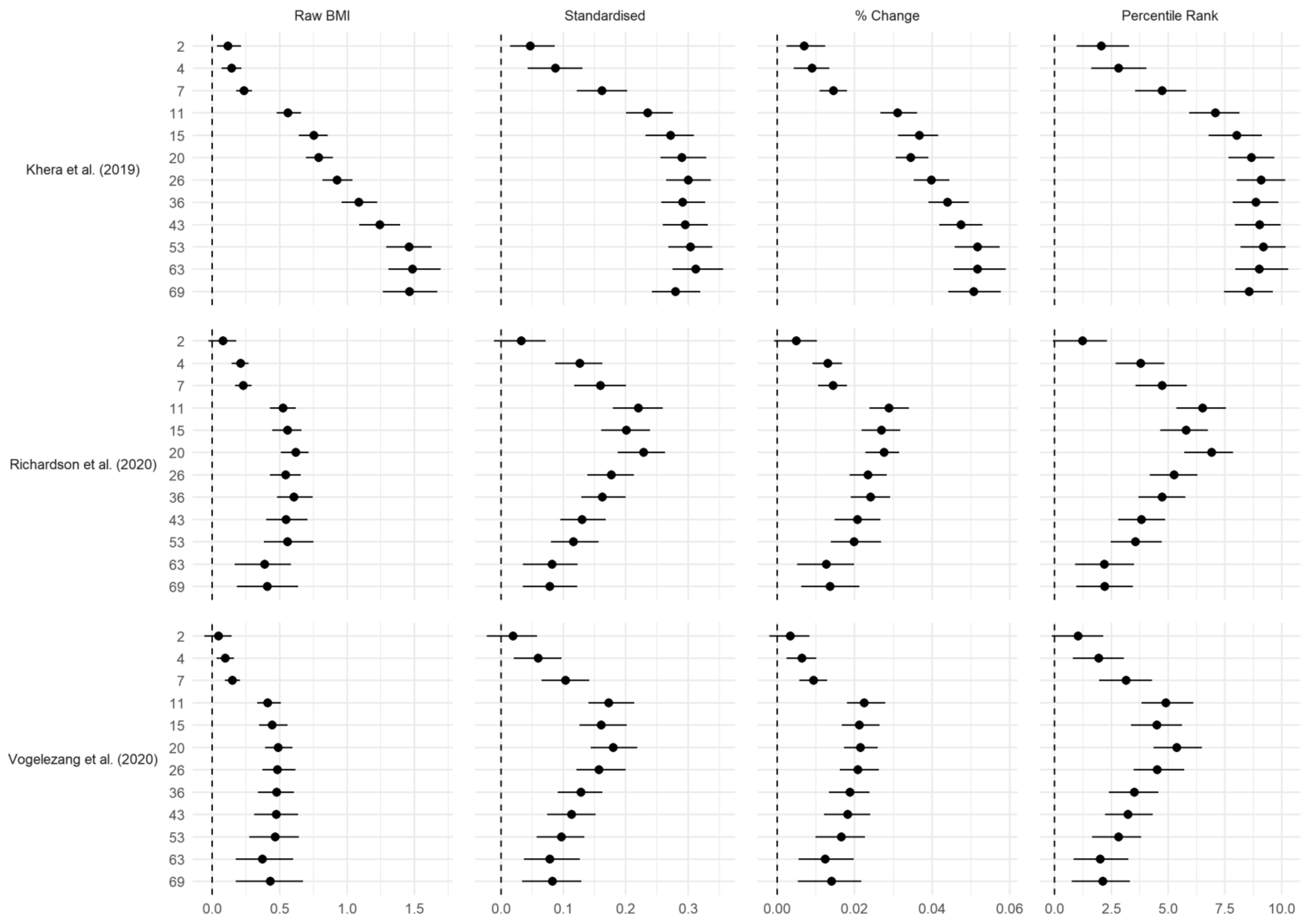


*Figure S2: Association between polygenic risk scores and BMI, measured as (left to right) raw scores, standardized values, logarithms, and percentile ranks. Standardization and percentile ranks calculated at each age at follow-up. Drawn from OLS regressions including adjustment for sex and repeated for each polygenic risk score and age at follow up. Confidence intervals estimated using bootstrapping (500 replications).*

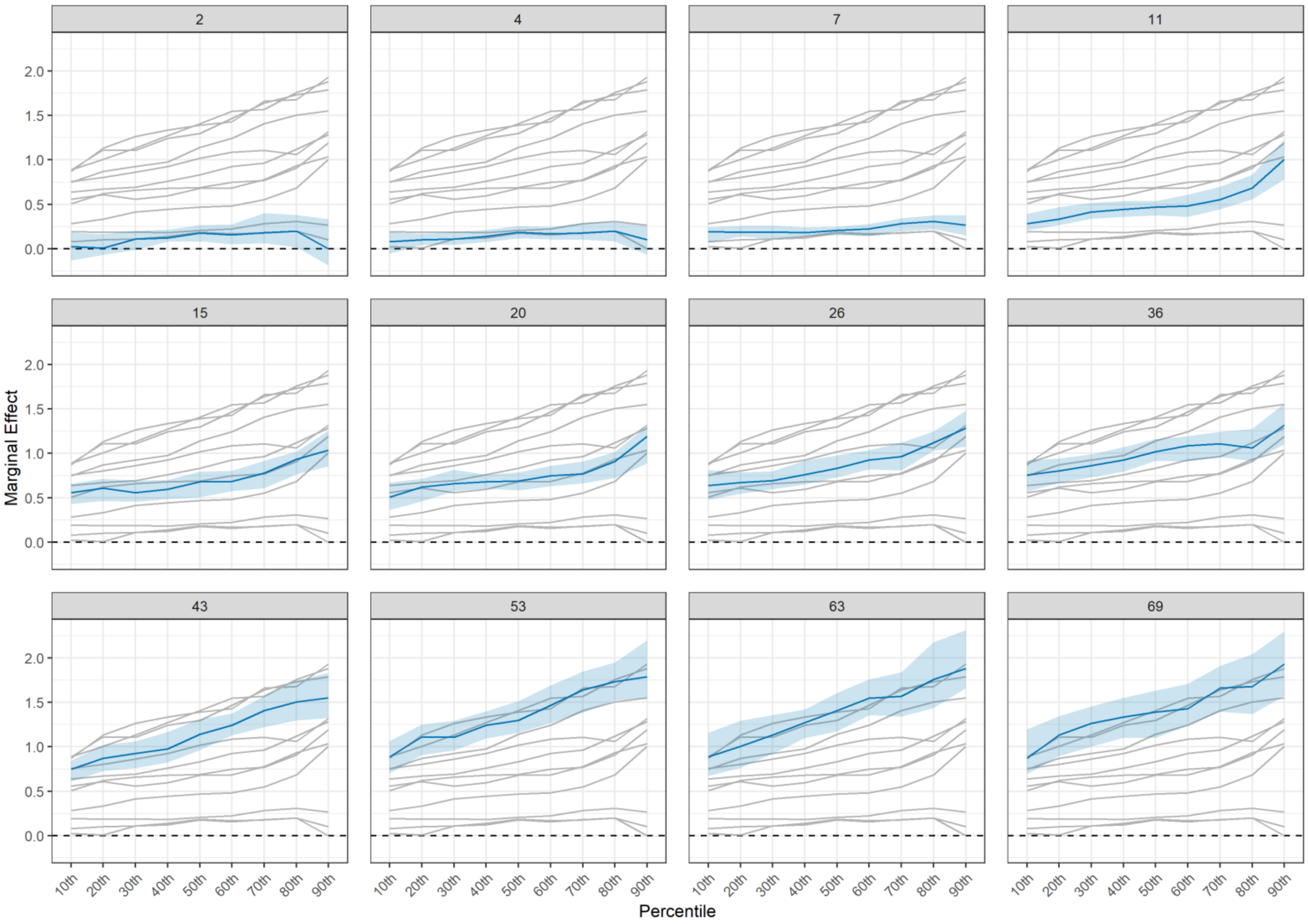


*Figure S3: Association between Khera et al. (2019) polygenic risk scores and (absolute) BMI. Drawn from quantile regressions including adjustment for sex, repeated at each follow up (panels) and decile (x-axis).*

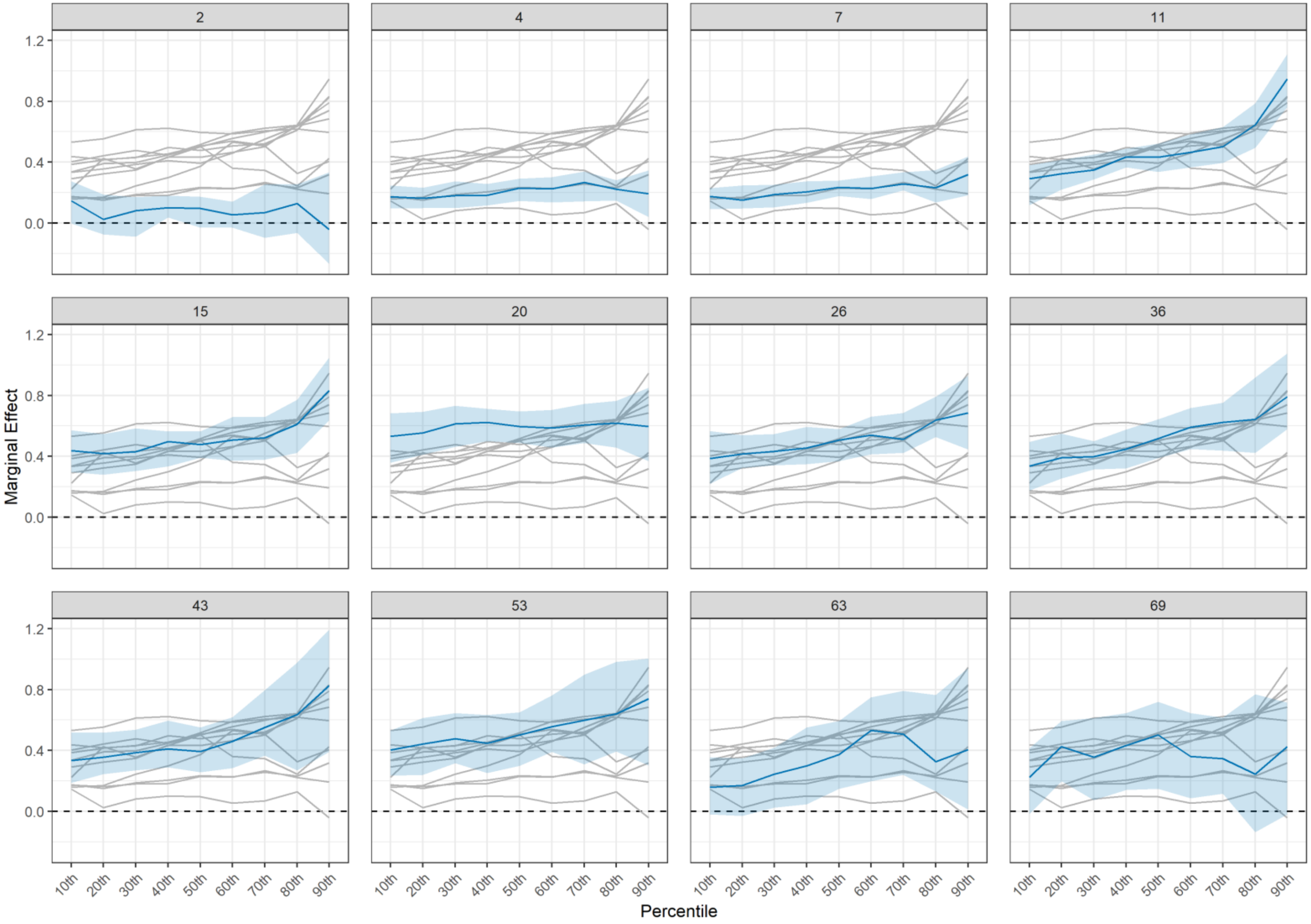


*Figure S4: Association between Richardson et al. (2020) polygenic risk scores and (absolute) BMI. Drawn from quantile regressions including adjustment for sex, repeated at each follow up (panels) and decile (x-axis).*

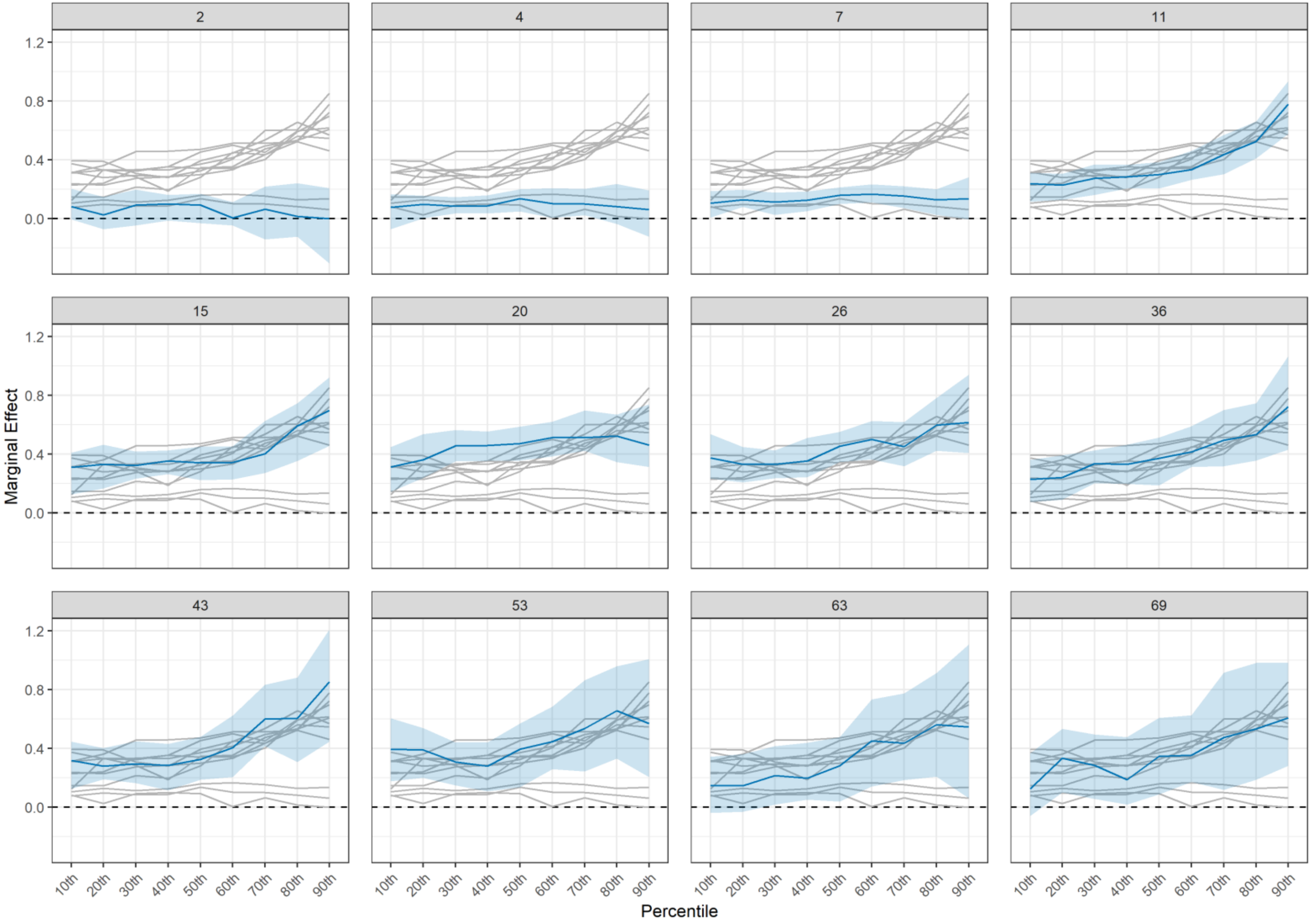


*Figure S5: Association between Vogelezang et al. (2020) polygenic risk scores and (absolute) BMI. Drawn from quantile regressions including adjustment for sex, repeated at each follow up (panels) and decile (x-axis).*

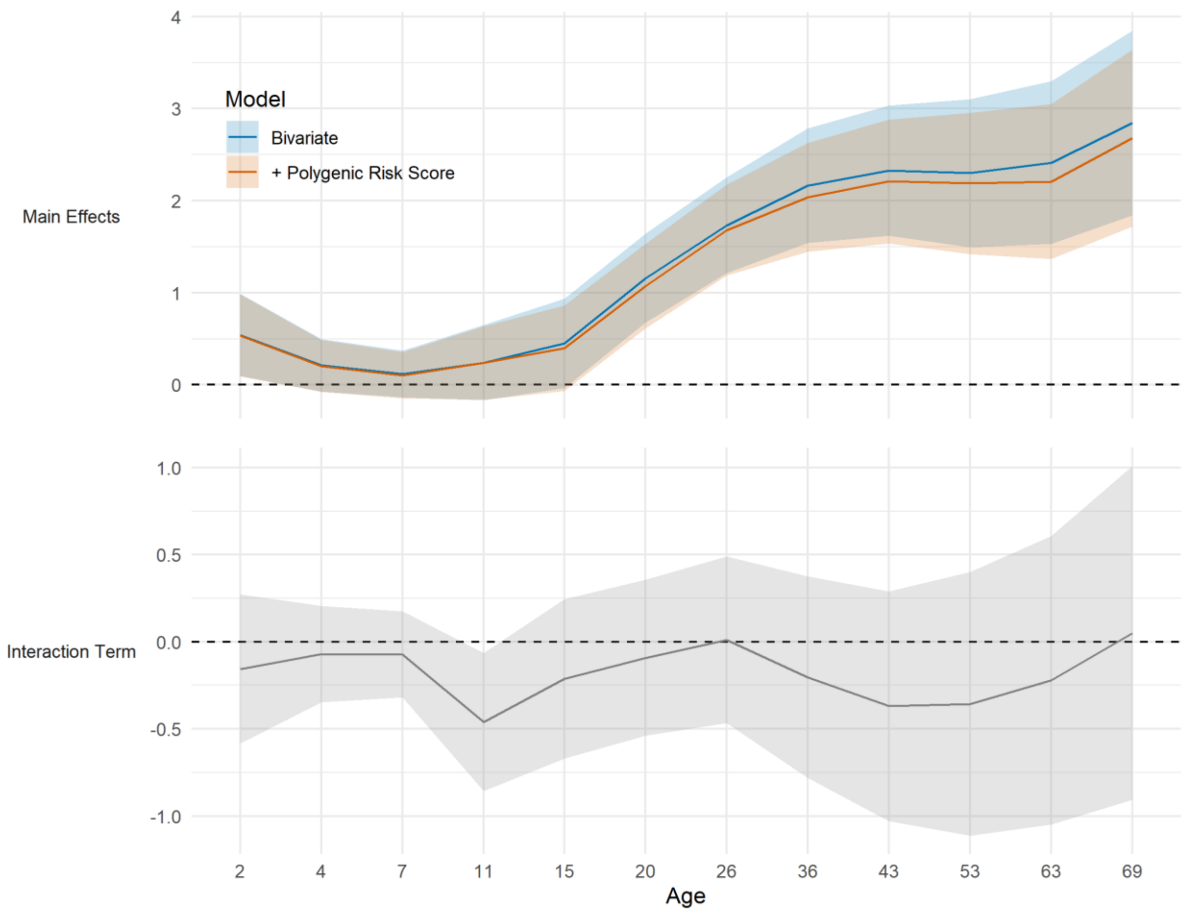


*Figure S6: Association between childhood socioeconomic position and body mass index (BMI) across life. Top panel shows the kg/m² difference in BMI in the lowest compared with highest socioeconomic position, before and after adjustment for Khera et al. (2019) polygenic index for higher BMI. Bottom panel shows coefficients for the social class x polygenic index interaction term (null line is evidence for no interaction). SEP measured as mother's education level converted to ridit score. Results from top panel drawn from OLS regression models including adjustment for sex (blue solid line) and further adjustment for polygenic indices (orange dashed line). Results from bottom panel drawn from OLS regression models including adjusted for sex, polygenic index score (Khera et al., 2019) and SEP.*

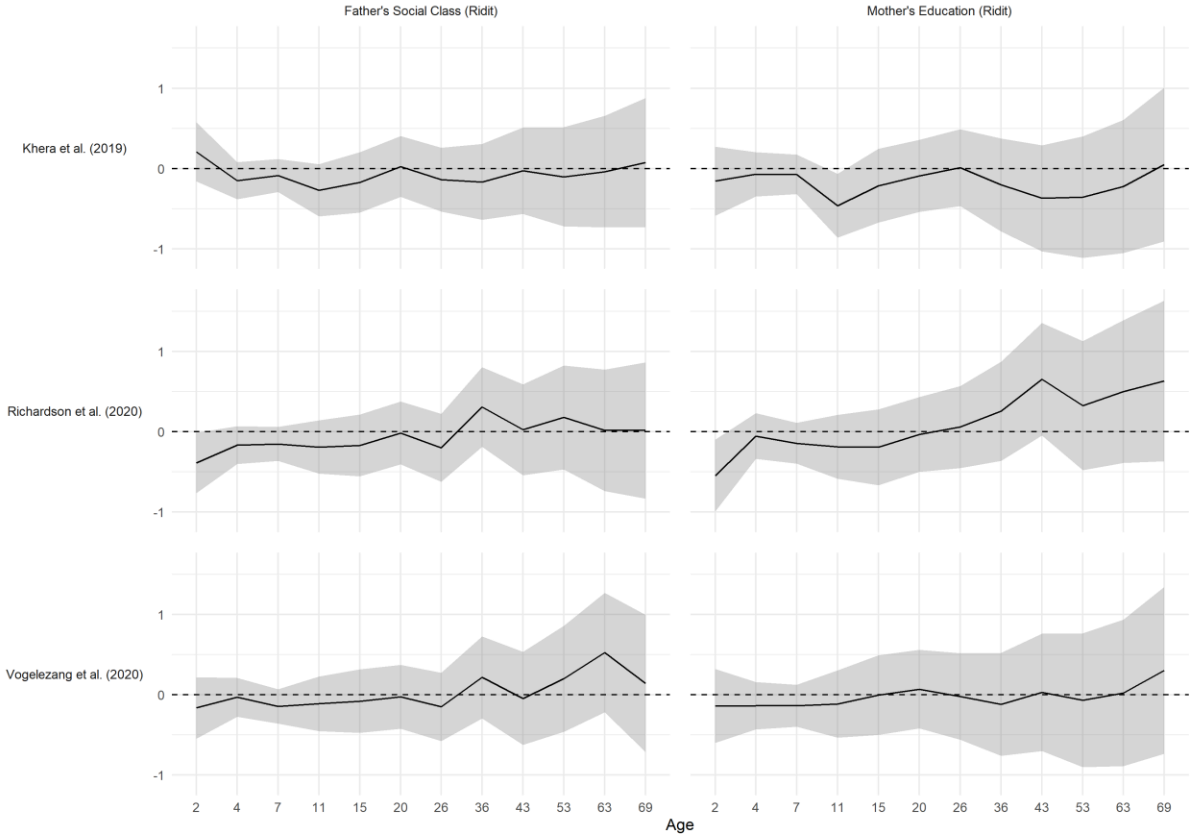


*Figure S7: Interaction effect between polygenic risk and childhood socioeconomic position; outcome is body mass index at each age (x-axis). Null line is evidence for no interaction. Results drawn from OLS regression models repeated for each definition of childhood SEP (columns) and polygenic index (rows) and including adjustment for sex, polygenic index score and SEP.*

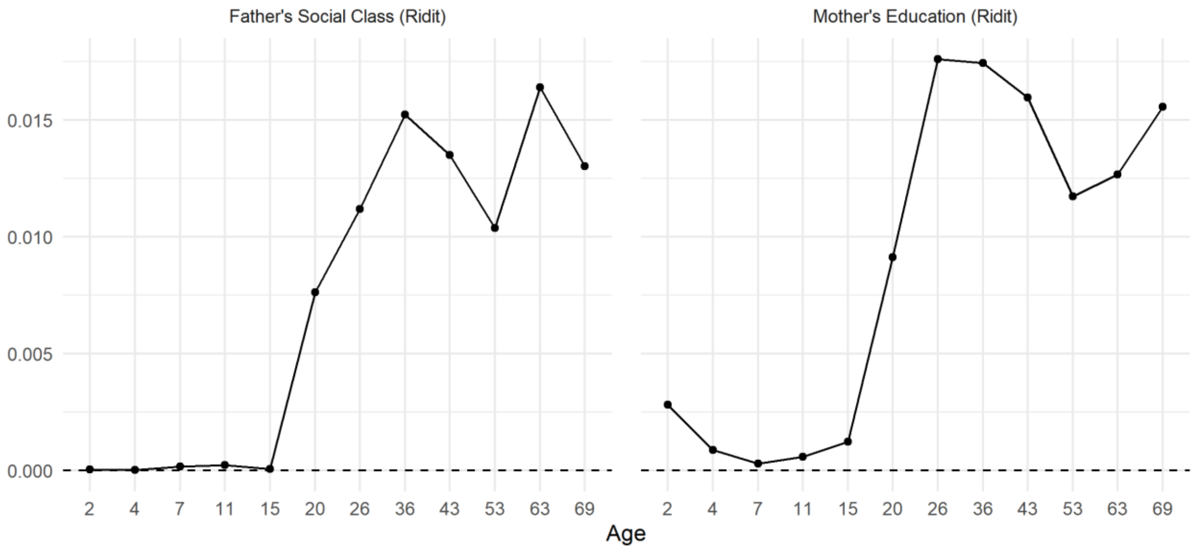


*Figure S8: Proportion of variation in BMI explained by childhood social position. Incremental $R^2$ compared to OLS regression model of BMI on sex and Khera et al. polygenic index. Left column: SEP measured as father's occupational class converted to ridit score. Right column: SEP measured as mother's education level converted to ridit score.*

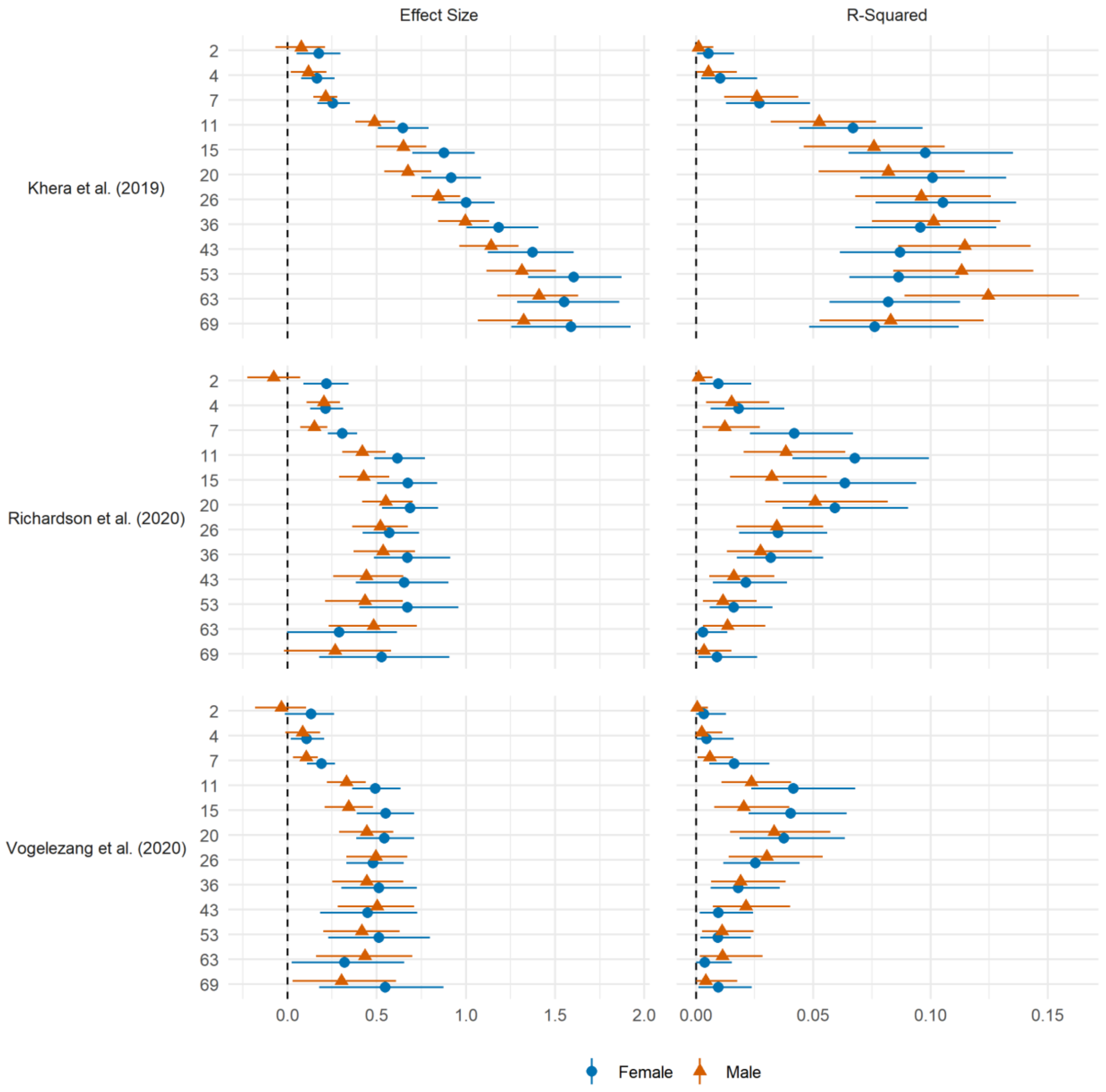


*Figure S9: Association between polygenic risk scores and BMI. Drawn from bivariate OLS regressions, repeated for each sex, polygenic risk score, and age at follow up. Confidence intervals estimated using bootstrapping (500 replications). Left panel: coefficient. Right panel: model* $R^2$.

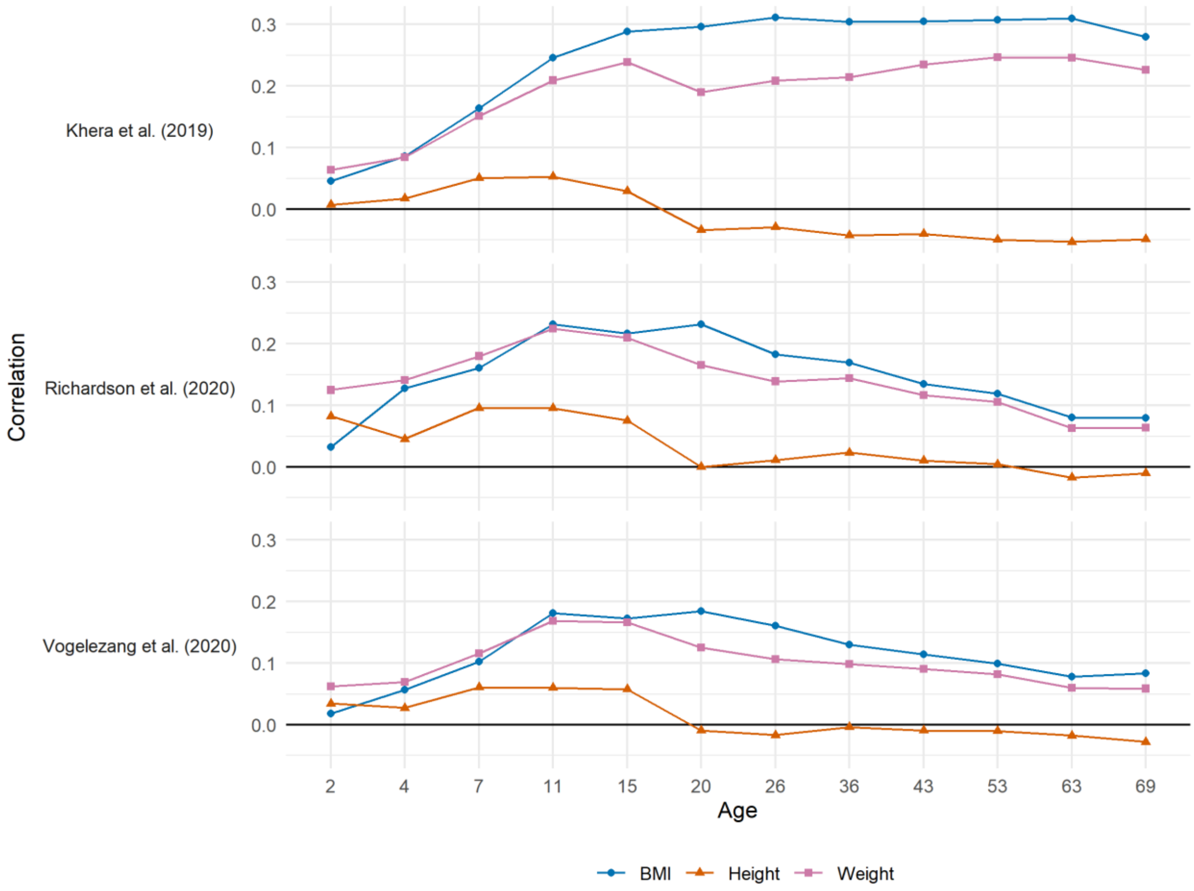


*Figure S10: Correlation by polygenic score and BMI, height and weight at age follow-up.*

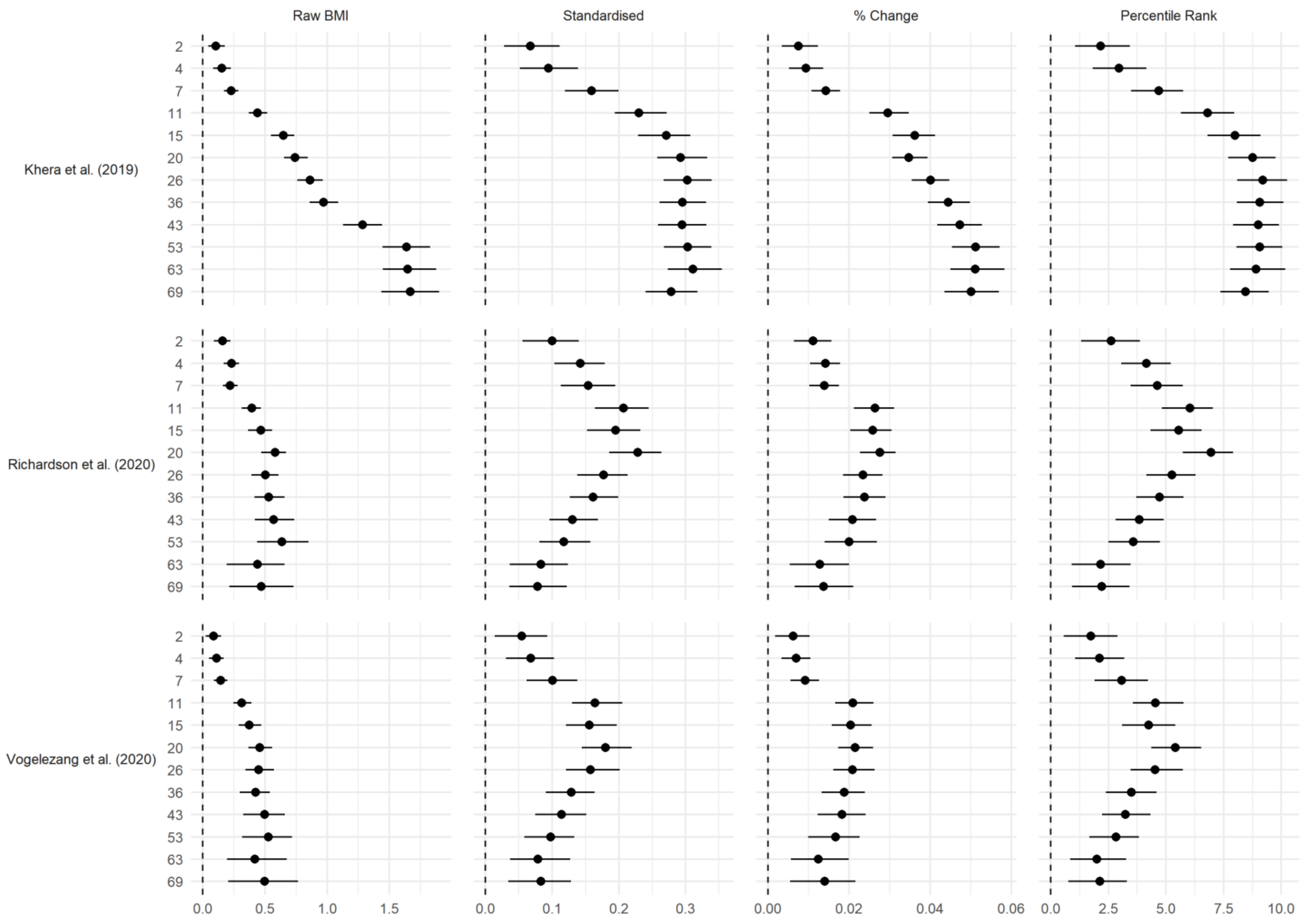


*Figure S11: Association between polygenic risk scores and corrected BMI, measured as (left to right) raw scores, standardized values, logarithms, and percentile ranks. Standardization and percentile ranks calculated at each age at follow-up. Correction calculated by finding x such that correlation between corrected BMI (kg/m$^x$) and height is zero at a given age. Drawn from OLS regressions including adjustment for sex and repeated for each polygenic risk score and age at follow up. Confidence intervals estimated using bootstrapping (500 replications).*

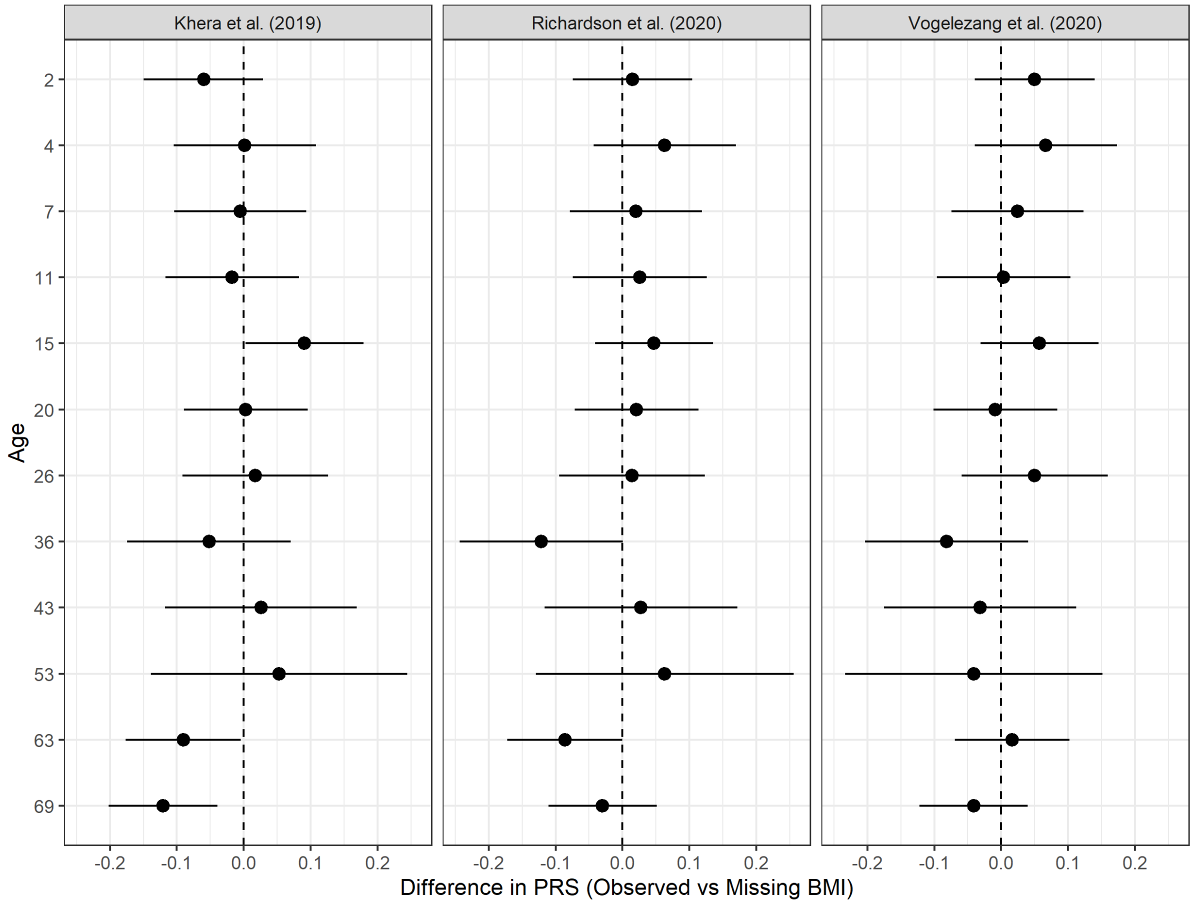


*Figure S12: Difference in average PRS scores (+ 95% CI) by whether participant had observed or missing BMI scores at a given age. Drawn from separate regressions for each combination of PRS score (columns) and age of follow-up (rows).*

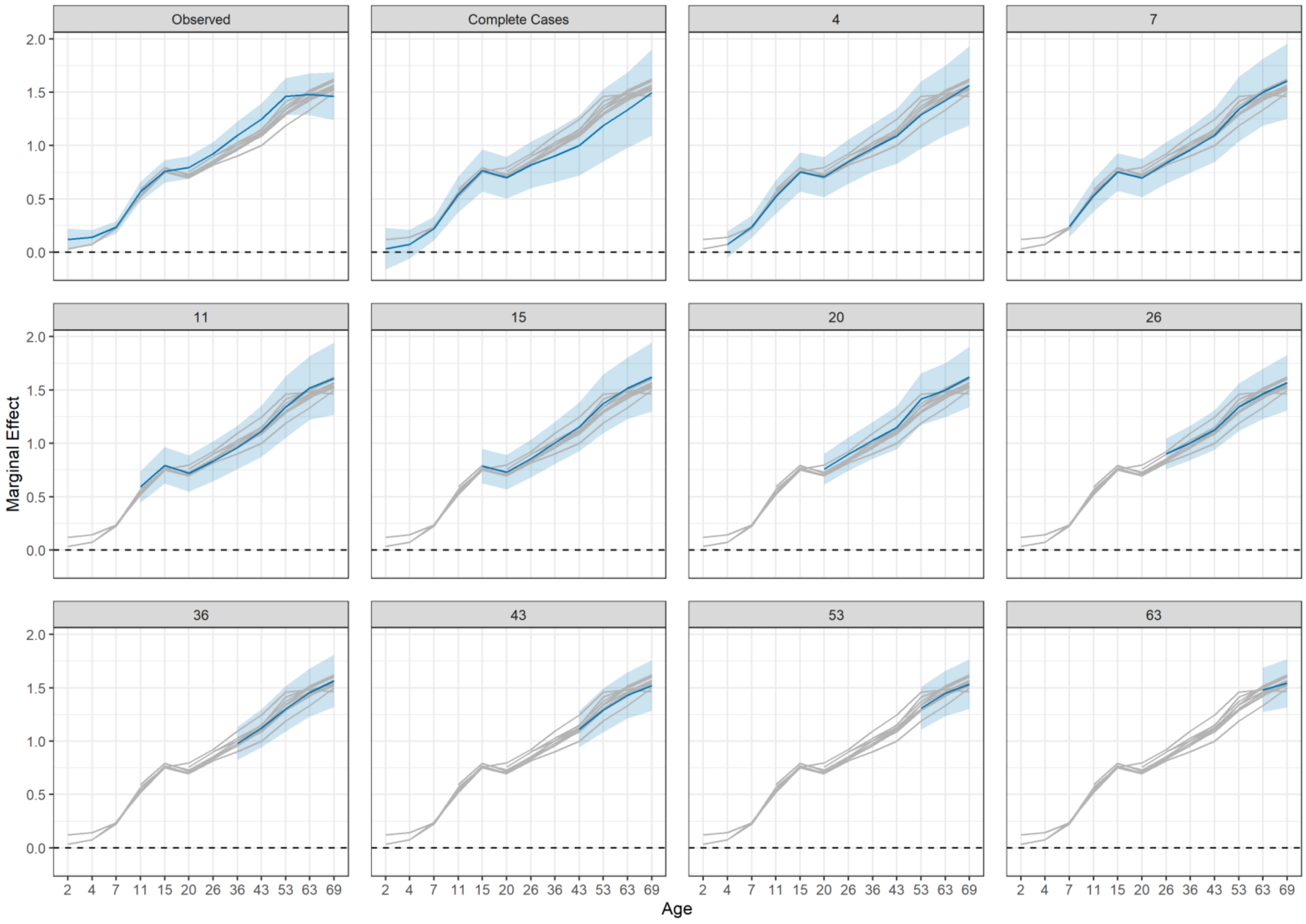


*Figure S13: Association between Khera et al. (2019) polygenic risk scores and (absolute) BMI. Drawn from OLS regressions including adjustment for sex and repeated at each follow up using observed sample and balanced samples of participants interviewed at each sweep following a given age (panels).*

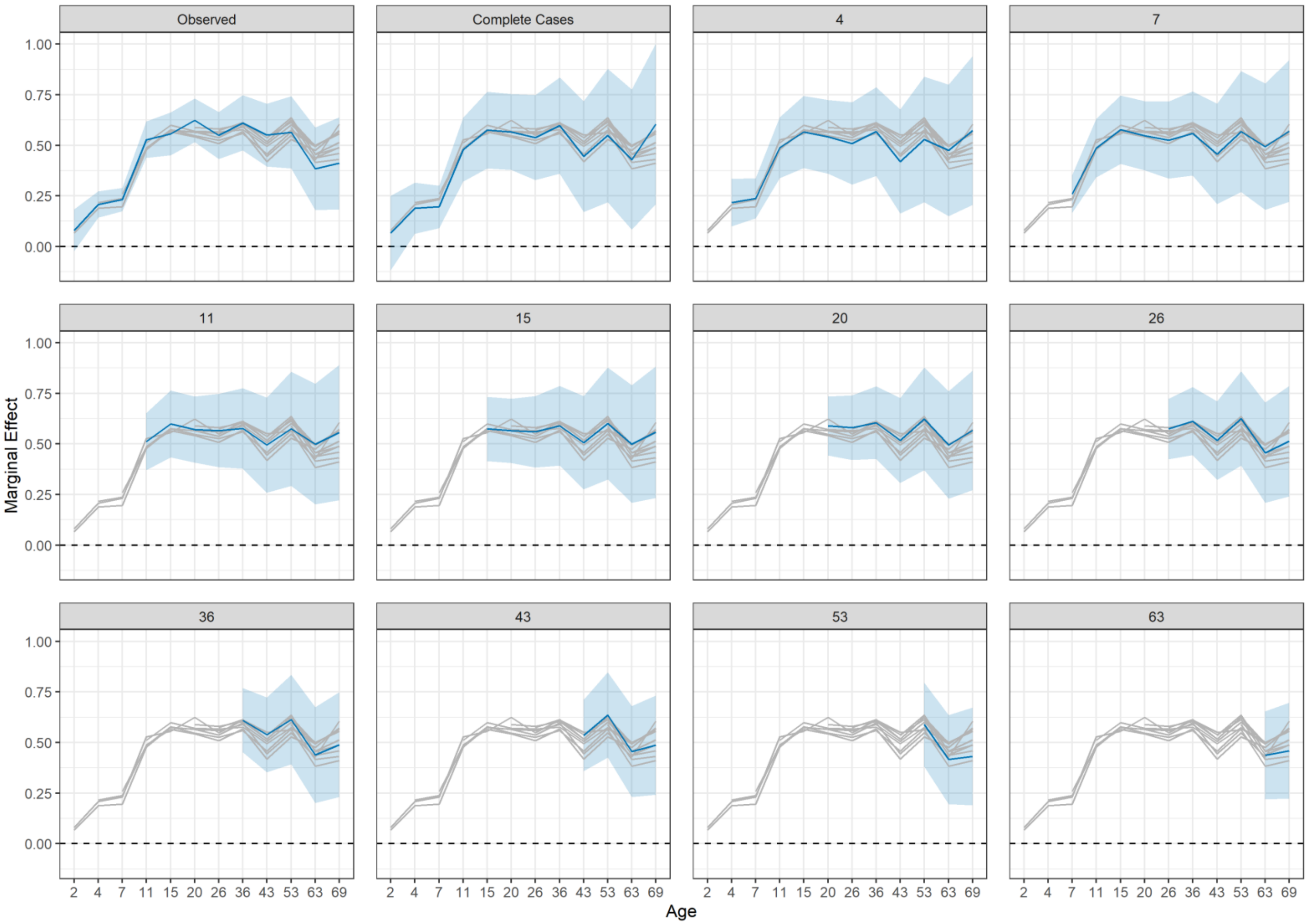


*Figure S14: Association between Richardson et al. (2020) polygenic risk scores and (absolute) BMI. Drawn from OLS regressions including adjustment for sex and repeated at each follow up using observed sample and balanced samples of participants interviewed at each sweep following a given age (panels).*

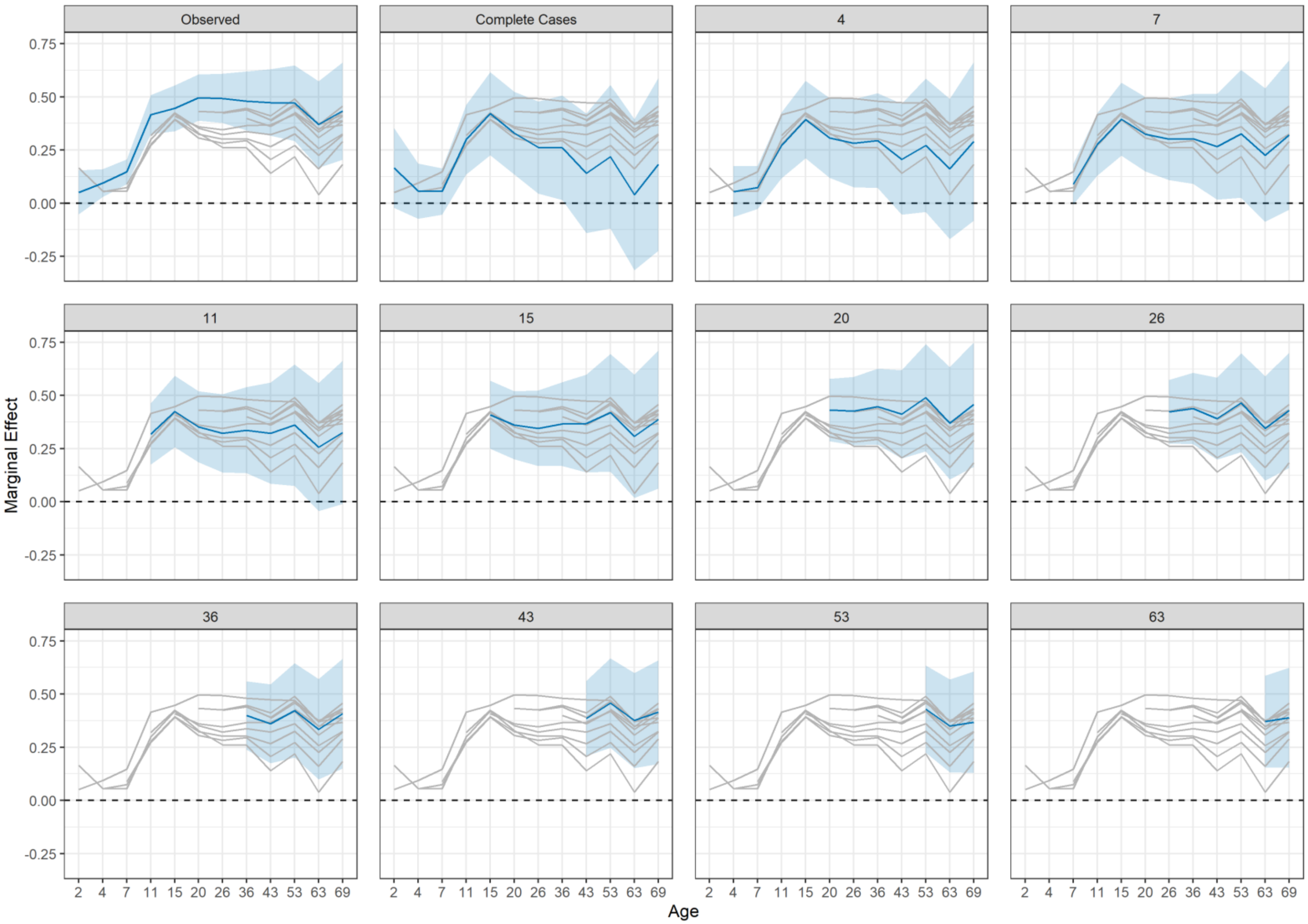


*Figure S15: Association between Vogelezang et al. (2020) polygenic risk scores and (absolute) BMI. Drawn from OLS regressions including adjustment for sex and repeated at each follow up using observed sample and balanced samples of participants interviewed at each sweep following a given age (panels).*

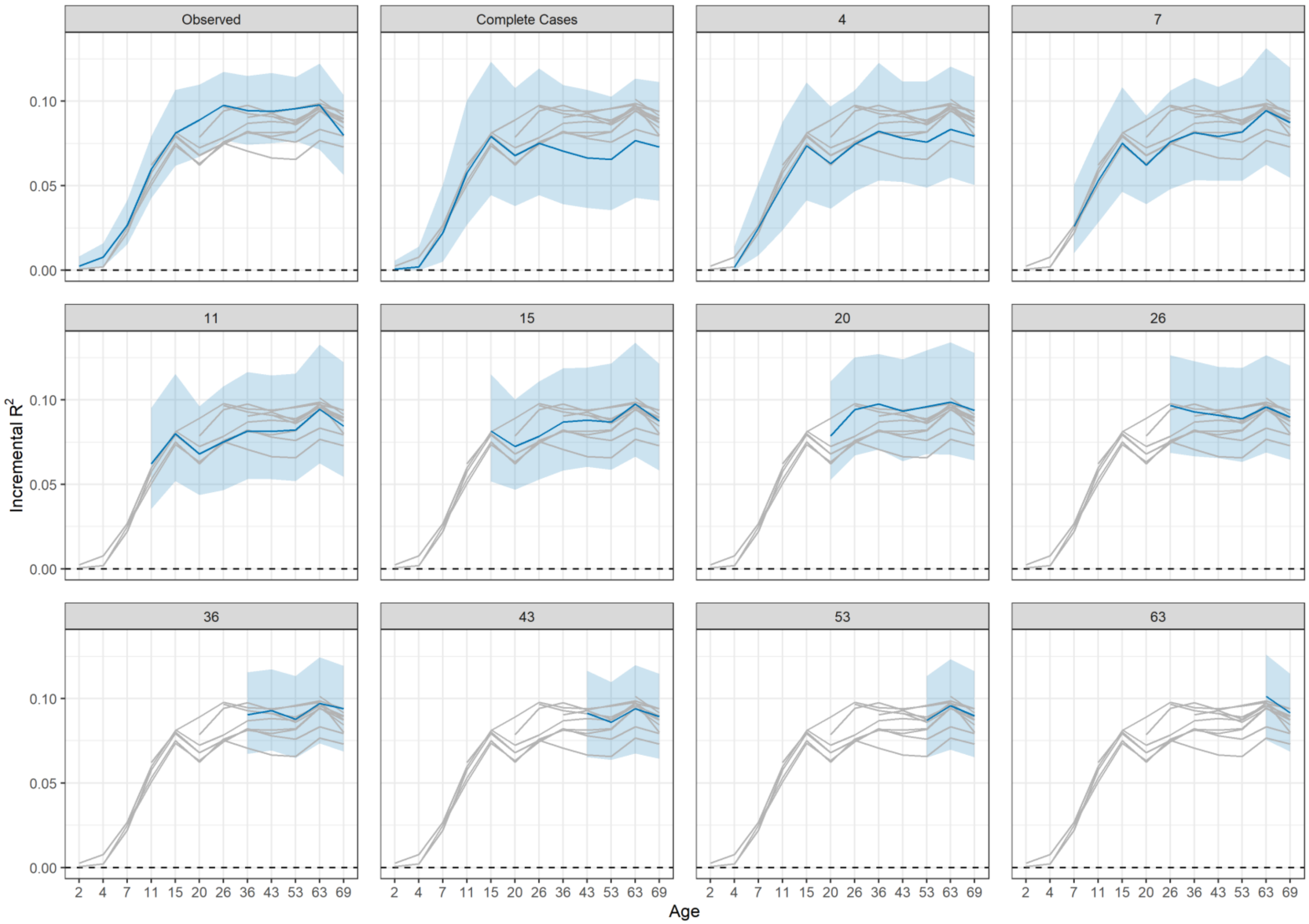


*Figure S16: Incremental proportion of variance in (absolute) BMI explained by Khera et al. (2019) polygenic risk score. Drawn from OLS regressions compared solely adjusting for sex. Regression models repeated at each follow up using observed sample and balanced samples of participants interviewed at each sweep following a given age (panels). Confidence intervals derived from 500 bootstrap replications using the centile method.*

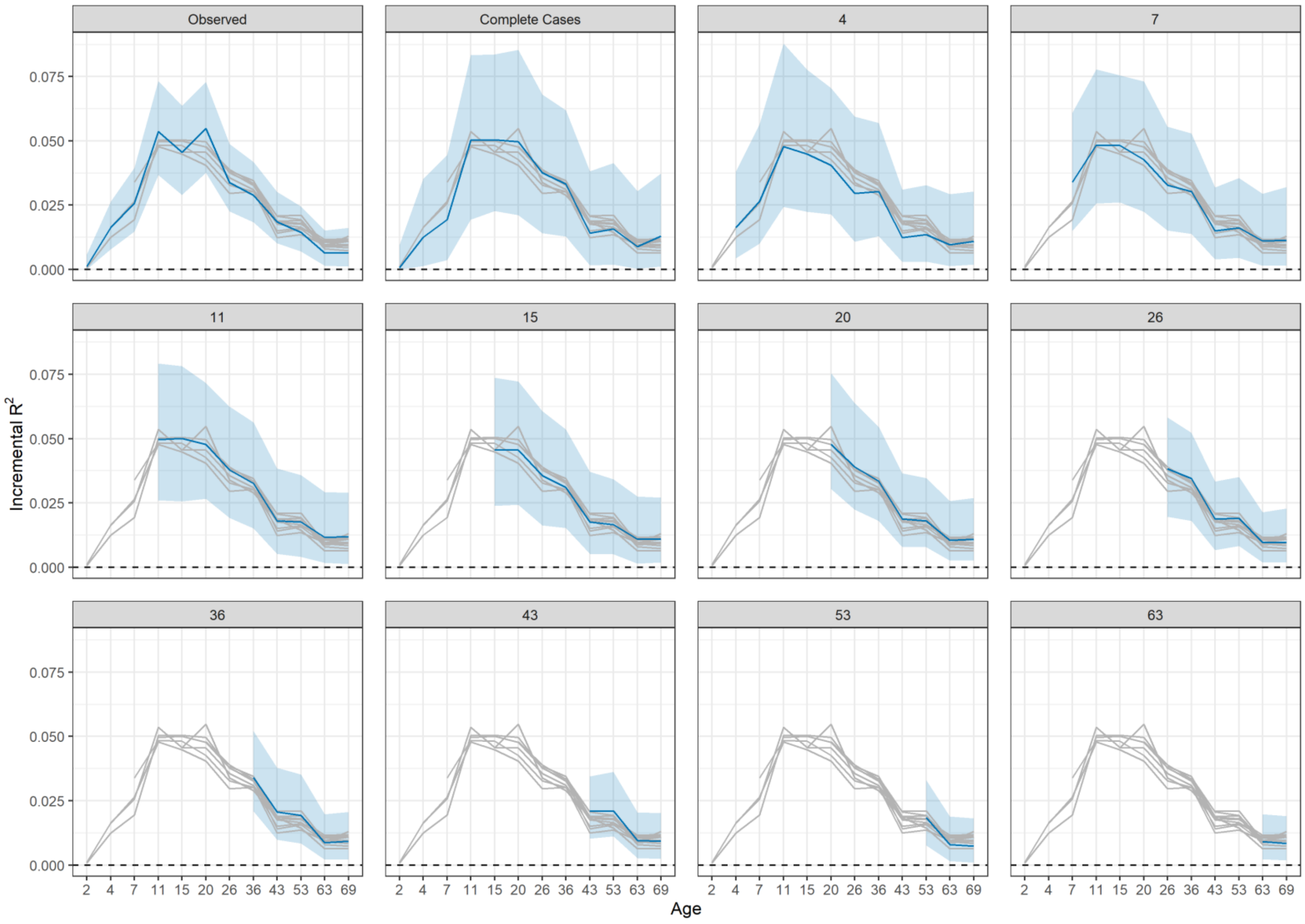


*Figure S17: Incremental proportion of variance in (absolute) BMI explained by Richardson et al. (2020) polygenic risk score. Drawn from OLS regressions compared solely adjusting for sex. Regression models repeated at each follow up using observed sample and balanced samples of participants interviewed at each sweep following a given age (panels). Confidence intervals derived from 500 bootstrap replications using the centile method.*

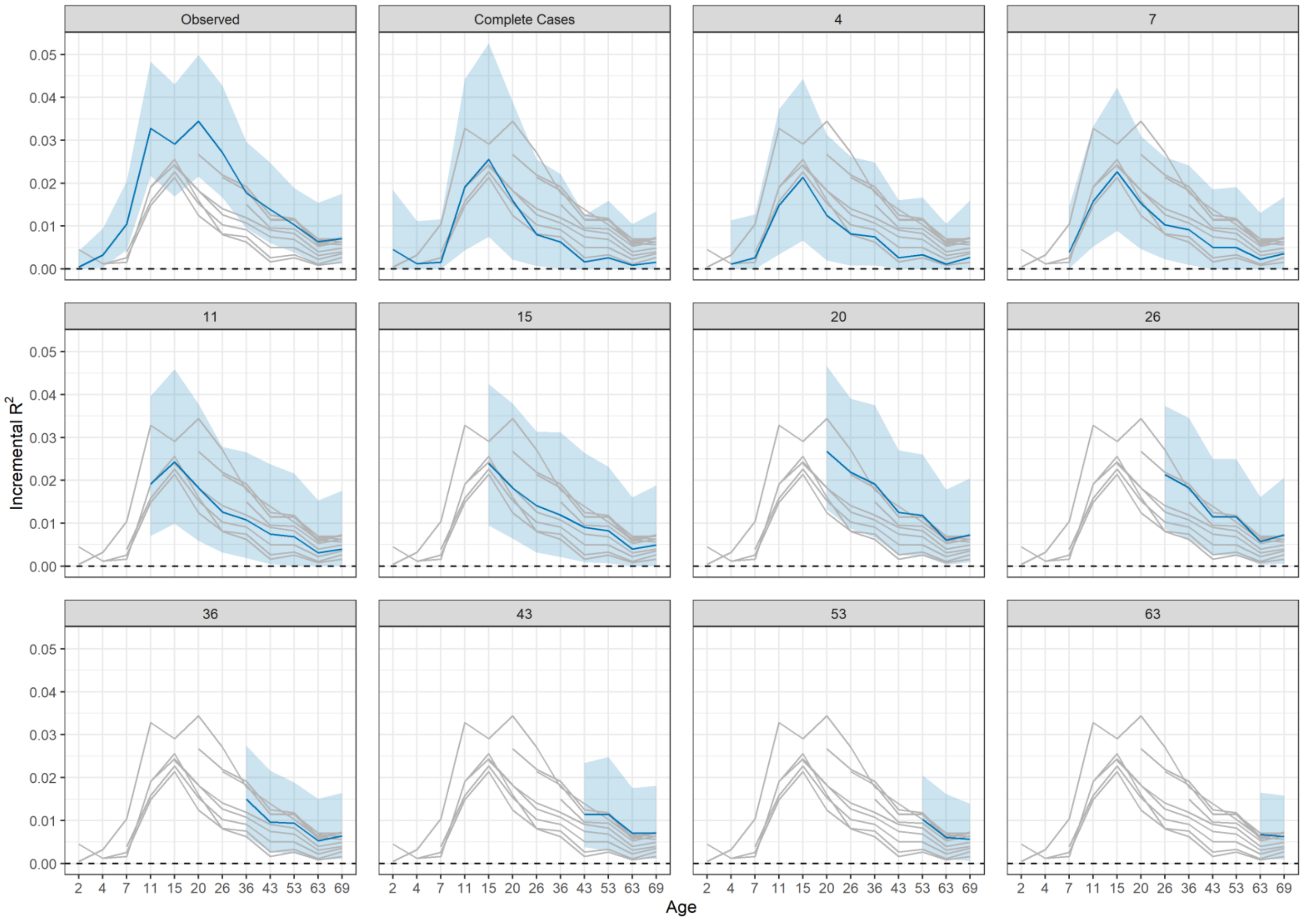


*Figure S18: Incremental proportion of variance in (absolute) BMI explained by Vogelezang et al. (2020) polygenic risk score. Drawn from OLS regressions compared solely adjusting for sex. Regression models repeated at each follow up using observed sample and balanced samples of participants interviewed at each sweep following a given age (panels). Confidence intervals derived from 500 bootstrap replications using the centile method.*

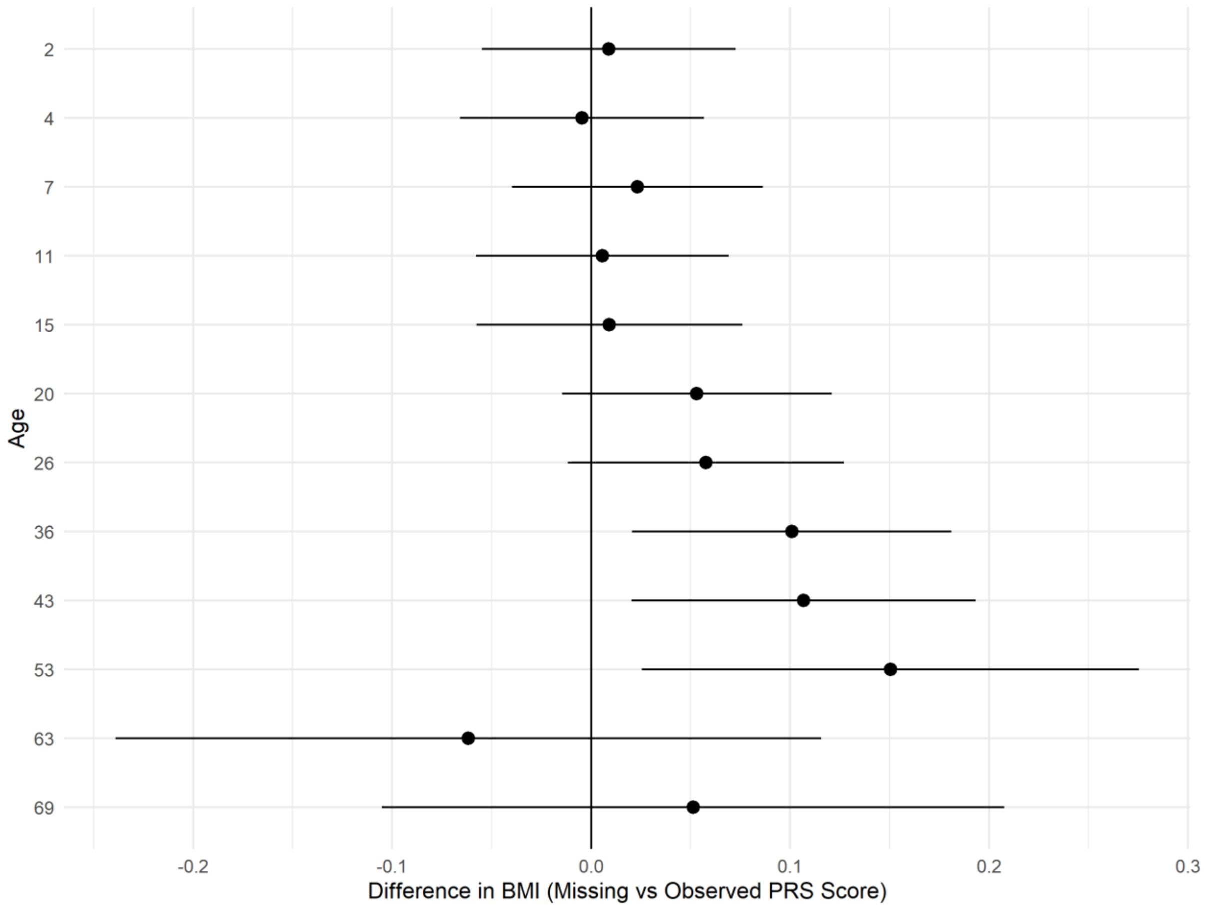


*Figure S19: Association between BMI and missing PRS score by age at follow-up (+ 95% confidence intervals). Drawn from linear regression models repeated at each age.*
